# Identifying and measuring important outcomes for evaluating the impact of pharmacist prescribing in Ireland: A modified Delphi study

**DOI:** 10.1101/2025.06.30.25330556

**Authors:** Ahmed Hassan Ali, Barbara Clyne, Judith Strawbridge, Melody Buckley, Mandy Daly, Michelle Flood, Siobhán Freeney, Caroline McCarthy, Frank Moriarty

**Author notes:** **Corresponding author name and address:** Frank Moriarty, School of Pharmacy and Biomolecular Sciences, Royal College of Surgeons in Ireland (RCSI) University of Medicine and Health Sciences, Dublin 2, Dublin, Ireland., **Corresponding author email address:**. **Funding** This research was funded by the Department of Health.

## Abstract

**Background:** Pharmacist role expansion, including pharmacist prescribing, is increasing globally. Assessing the impact of such role expansion is vital.

**Purpose:** To identify key outcomes to evaluate pharmacist prescribing within Ireland’s planned Common Conditions Service (CCS) and independent pharmacist prescribing contexts, and determine their measurability using existing health data sources.

**Methods:** In a three-round Delphi study, an expert panel (including pharmacists, pharmacy technicians, prescribers, public/patient contributors, policymakers, academics) rated the importance of outcomes, pre-identified via a rapid overview of reviews, using Likert scales. The research team involved public/patient team members throughout the study. Outcomes reaching consensus for inclusion (rated by ≥75% as critically important) or exclusion (<25%) after the second round were not re-rated. In the third round, remaining outcomes were re-rated and experts also rated the feasibility of measuring each outcome using existing health data sources.

**Results:** Thirty experts completed all rounds. For CCS, seven outcomes reached consensus as critically important: Patient experience and satisfaction, Access to care, Guideline concordance, Symptom improvement, or clinical cure, Re-consultation with other health care providers, Cost of care to the healthcare system, and Referral to other healthcare providers. For other independent prescribing contexts, ten outcomes were critically important, including Mortality, Clinical effectiveness, Adverse events, and Cost of care to patients. Important CCS outcomes varied in their measurability using existing data, with Prescribing rate (76% agreement) and Cost to patients (64% agreement) were most feasible.

**Conclusions:** This study identified priority outcomes for evaluating the impact of pharmacist prescribing, encompassing clinical, safety, economic, and patient perspectives.

**Highlights:**

- Clinical, economic, safety, and patient-reported outcomes are key for assessing the impact of pharmacist prescribing.
- Seven priority outcomes were identified for Common Conditions Services, and ten for wider independent prescribing contexts.
- Existing health data can support outcome measurement, but enhancements are needed to improve interoperability and coverage.

## Background

Healthcare systems worldwide are facing a critical shortage of skilled professionals, with a needs-based global shortfall projected to exceed ten million by 2030.^1, 2, 3^ This growing gap is driven by increasing demand for primary care services due to population growth, an ageing demographic, longer life expectancy, and a higher prevalence of comorbidities.^1, 2^ Additionally, the expansion of disease definitions encompassing pre-disease populations and associated growth in preventive care over recent decades can also exacerbate the shortfall in supply in high-income systems.^4^ Structural workforce challenges including understaffing, excessive workload, and adverse work conditions also compound the growing service pressures, undermining staff well-being, workforce retention and patient outcomes.^5, 6^ The shortage of healthcare providers, particularly in primary care, has led to challenges in accessing timely care and has placed increasing pressure on emergency departments.^1, 2, 7^

In response, countries such as the United Kingdom, Canada, the United States, Australia, New Zealand, and France have introduced strategies that expand the roles of healthcare professionals such as pharmacists by incorporating task-shifting, which involves transferring selected clinical activities, such as prescribing, from physicians to other appropriately trained healthcare professionals, to improve access to health care.^1, 2, 7^ Within the Irish context, the evolving healthcare landscape involving Sláintecare health reforms (aiming to deliver integrated, high-quality, efficient, accessible, and cost-effective care close to the patients) has led to proposals for introducing pharmacist prescribing to ease the burden on general practitioners and enhance timely management of common conditions.^8, 9^ In Ireland, an Expert Taskforce was established in 2023 to support the expansion of the role of pharmacists.^10^ Their final report recommended that pharmacists be “enabled to exercise independent, autonomous prescriptive authority within and related to the individual practitioner’s scope of practice and competence”. It recommends focusing initially on prescribing for common conditions within community pharmacy, enabling pharmacists to manage common conditions by offering advice, and, when appropriate, prescribing prescription-only medicines through established protocols for an initial list of common conditions such as allergic rhinitis, cold sores, conjunctivitis, impetigo, oral thrush, shingles, uncomplicated urinary tract infections (UTIs)/cystitis, and vulvovaginal thrush.^8^ The report recommends then progressing to full prescribing authority through implementing models of pharmacist prescribing within primary and secondary care settings.

The expansion of pharmacists’ roles presents a significant opportunity to enhance the delivery of high-quality, accessible, and cost-effective healthcare services.^8^ In addition, it facilitates greater utilisation of pharmacists’ clinical expertise relating to medications and may support workforce retention by addressing factors such as career fulfilment, professional development, and recognition of their contributions.^11^ As pharmacists’ roles are expanded, assessing their impact is crucial to ensure they enhance patient care and health outcomes. The efficiency of such evaluations can be enhanced substantially by measuring relevant outcomes from routinely collected data. Compared to collecting data or establishing datasets for specific research or evaluation aims, routine data captured within existing healthcare data/systems can reduce costs, time, and other resources. Such data can be used for service evaluation and/or research in various ways e.g. evaluating the introduction of prescribing on overall quality of antimicrobial prescribing via an interrupted time series study. Therefore, it is important to understand the outcomes that should be collected to evaluate the impact of these new roles, and the potential to measure these outcomes via routinely collected data.

This study aimed to identify key outcomes that should be measured to assess the impact of introducing pharmacist prescribing, both generally, and more specifically related to a Common Conditions Service (CCS), using a Delphi approach with a diverse panel of experts that included healthcare professionals, policymakers, academic experts, and members of the public. A secondary aim was to assess how existing health datasets in the Irish setting could be used to optimise data collection for these outcomes in the context of a CCS.

## Methods

This study used a Delphi approach to achieve consensus among a panel of experts on important outcomes with data collected between January and March 2025. In addition to rating outcome importance, an expanded expert panel (i.e. with additional experts in health data sources and systems) were asked to consider the feasibility of measuring these outcomes using existing health datasets and sources. The Delphi process was limited to three rounds as predefined in the protocol. This Delphi study formed part of a broader multi-phase outcome prioritisation process, starting with a rapid overview of existing reviews to identify relevant outcomes. This approach aligns with established procedures for the development of Core Outcome Sets (COS) as outlined by the Core Outcome Measures in Effectiveness Trials (COMET) Initiative. This typically involves evidence synthesis such as systematic or rapid review, followed by a consensus-based approach like a Delphi study.^12^ Ethical approval was granted by the RCSI University of Medicine and Health Sciences Research Ethics Committee (REC) (reference number REC202411038). The study is reported in accordance with the DELPHISTAR reporting guidelines and the GRIPP2 short form for Patient and Public Involvement (PPI).^13, 14^

### Participants

Non-probability sampling (purposive and snowball) was used to recruit the Delphi panel, aiming to involve 20-25 experts (in line with previous research) based on their expertise and/or experience in a relevant sector,^15, 16, 17^ with a target of 4–5 experts from each of the following groups:

- Pharmacists and pharmacy technicians from a range of settings (e.g. primary and secondary care settings)
- Prescribers from different settings and professions (doctors, nurses, dentists)
- Patients/members of the public
- Those working in relevant policy, regulatory or other statutory public bodies, and
- Academics, in particular those with expertise in pharmacist prescribing and outcome measurement.

Experts were invited via targeted invitations to individuals from the professional networks of the project team, including those with experience as patient and public involvement (PPI) contributors in research and/or policy. Invitations outlined that the study would involve completing three online questionnaires, each requiring approximately 30 minutes. Recipients were asked to share the invitation with a colleague with relevant expertise and/or experience if they were unable to participate themselves. The invitation contained a participant information leaflet and a link to a Microsoft Forms (Microsoft Corporation, Redmond, WA) questionnaire containing an electronic consent form followed by the first-round questions. In addition to the experts recruited from the first round for the full Delphi process, additional experts in health data sources and systems in Ireland were invited in the third round. These participants were recruited through the same process as the initial panel, forming an expanded expert panel to answer the additional questions on how to measure important outcomes.

### First round

A preliminary list of outcomes was identified from a rapid overview of reviews. This involved systematically identifying, synthesising, and extracting outcomes previously reported in evidence syntheses related to the evaluation of pharmacist prescribing.^18^ The identified outcomes were reviewed, adapted, and refined by the research team members to ensure relevance and clarity (see Appendix 1), resulting in 18 outcomes for prescribing in CCS and 15 for other independent prescribing.

The first-round survey consisted of two sections, reflecting the main phases of the proposed introduction of pharmacist prescribing (CCS and other independent prescribing), and Delphi experts were asked to rate their agreement with the importance of each outcome (Table 1, with some modifications made by the research team to separate distinct outcomes as in Appendix 1). This was assessed on a 9-point Likert scale, labelled as follows: 1–3, not important; 4–6 important but not critical; 7–9, critically important. A brief description of the outcome accompanied each question, providing additional details to ensure consistency in interpretation (see Appendix 2). Panel members were also asked to provide rationale for their ratings or any suggestions for modifications to outcomes in free-text boxes following each outcome, or suggest additional outcomes that should be included in free-text boxes at the end of each section. For illustrative purposes, a sample of the first-round questionnaire is presented in Appendix 3.

**Table 1.** Overall results of ratings of importance of outcomes across rounds

| Outcomes | First round |  | Second round |  | Third round |  | Decision |
| --- | --- | --- | --- | --- | --- | --- | --- |
|  | Median | % rated<br>7-9 | Median | % rated<br>7-9 | Median | % rated<br>7-9 |  |
| Common Conditions Service |  |  |  |  |  |  |  |
| Symptom resolution or improvement, or clinical cure* | 7 | 68.6 | 7 | 78.8 |  |  | Critically Important<sup>†</sup> |
| Re-consultation with the pharmacist | 7 | 54.3 | 7 | 54.6 | 6.5 | 50.0 | Medium Priority<sup>††</sup> |
| Referral to other healthcare providers | 7 | 74.3 | 7 | 75.8 |  |  | Critically Important<sup>†</sup> |
| Re-consultation with other health care providers/other healthcare utilisation | 7 | 62.9 | 7 | 66.7 | 7 | 76.7 | Critically Important<sup>††</sup> |
| Prescribing rate | 6 | 48.6 | 7 | 51.5 | 6.5 | 50.0 | Medium Priority<sup>††</sup> |
| Guideline concordance/appropriateness of medications | 8 | 82.9 | 8 | 84.9 |  |  | Critically Important<sup>†</sup> |
| Patient adherence to medication | 5 | 25.7 | 5 | 12.1 |  |  | Low Priority<sup>†</sup> |
| Adverse events** | 7 | 57.1 | 7 | 57.6 | 7 | 70.0 | Medium Priority<sup>††</sup> |
| Patient experience and satisfaction with care | 8 | 85.7 | 8 | 90.9 |  |  | Critically Important<sup>†</sup> |
| Quality of life | 5 | 22.9 | 4 | 12.1 |  |  | Low Priority<sup>†</sup> |
| General health status*** | 4 | 5.7 |  |  |  |  |  |
|  | Median | % rated<br>7-9 | Median | % rated<br>7-9 | Median | % rated<br>7-9 |  |
| Patient functioning*** | 5 | 11.4 |  |  |  |  |  |
| Cost of care to patients | 7 | 60.0 | 7 | 57.6 | 7 | 63.3 | Medium<br>Priority<sup>++</sup> |
| Cost of care to providers | 7 | 57.1 | 6 | 48.5 | 6 | 23.3 | Low<br>Priority<sup>++</sup> |
| Cost of care to the<br>healthcare system | 7 | 71.4 | 7 | 75.8 |  |  | Critically<br>Important<sup>+</sup> |
| GP workload | 6 | 42.7 | 6 | 33.3 | 6 | 16.7 | Low<br>Priority<sup>++</sup> |
| Access to care | 7 | 74.3 | 7 | 87.9 |  |  | Critically<br>Important<sup>+</sup> |
| Level of service activity | 6 | 40.0 | 6 | 18.2 |  |  | Low<br>Priority<sup>+</sup> |
| Other Independent Prescribing |  |  |  |  |  |  |  |
| Mortality | 7 | 60.0 | 7 | 66.7 | 7 | 76.7 | Critically<br>Important<sup>++</sup> |
| Clinical effectiveness | 7 | 74.3 | 8 | 78.8 |  |  | Critically<br>Important<sup>+</sup> |
| Healthcare utilisation | 7 | 60.0 | 7 | 60.6 | 7 | 76.7 | Critically<br>Important<sup>++</sup> |
| Prescribing pattern (rates<br>and changes) | 7 | 51.4 | 7 | 54.6 | 7 | 66.7 | Medium<br>Priority<sup>++</sup> |
| Guideline concordance/<br>appropriateness of<br>medications | 7 | 82.9 | 7 | 90.9 |  |  | Critically<br>Important<sup>+</sup> |
| Patient adherence to<br>medication | 6 | 25.7 | 6 | 15.2 |  |  | Low<br>Priority<sup>+</sup> |
| Adverse events | 7 | 62.9 | 7 | 69.7 | 7 | 86.7 | Critically<br>Important<sup>++</sup> |
| Patient experience and<br>satisfaction with care | 7 | 74.3 | 7 | 84.9 |  |  | Critically<br>Important<sup>+</sup> |
| Quality of life | 6 | 37.1 | 6 | 33.3 | 6 | 33.3 | Medium<br>Priority<sup>++</sup> |
| General health status*** | 5 | 22.9 |  |  |  |  |  |
| Patient functioning*** | 6 | 25.7 |  |  |  |  |  |
| Cost of care to patients | 7 | 68.6 | 7 | 72.7 | 7 | 76.7 | Critically<br>Important<sup>++</sup> |
| Cost of care to service<br>providers | 7 | 60.0 | 7 | 69.7 | 7 | 90.0 | Critically<br>Important<sup>++</sup> |
|  | Median | % rated<br>7-9 | Median | % rated<br>7-9 | Median | % rated<br>7-9 |  |
| Cost of care to the healthcare system | 7 | 68.6 | 7 | 72.7 | 7 | 86.7 | **Critically Important<sup>††</sup>** |
| Access to care | 7 | 68.6 | 7 | 84.9 |  |  | **Critically Important<sup>†</sup>** |

\* Outcome was rephrased from “Clinical cure/symptom resolution or improvement” in the first round to “Symptom resolution or improvement, or clinical cure” in subsequent rounds based on the expert panel feedback.
\* \* Outcome was rephrased from “adverse effects” in the first round the first round to “adverse events” in subsequent rounds based on expert panel feedback.
\* \* \* Outcomes were merged and rated under “quality of life” after the first round, based on expert panel feedback and high correlation of ratings.
† Consensus was reached based on results of the second round.
†† Consensus was reached based on results of the third round.

### Second round

In the second round, Delphi experts re-evaluated the importance of the same outcomes assessed in the first round, with some modifications informed by expert feedback and a note of clarification that ratings should be based on importance, without consideration of the feasibility of measurement. Wording of some outcomes and their descriptions were also modified, and “Patient functioning” and “General health status” were incorporated into “Quality of life”. The ratings and the rationales provided in the first round were summarised, both for and against the importance of the outcome. Summaries of the distributions of the ratings across the panel (i.e. bar chart of responses) were embedded in the questionnaire, with a synthesis of rationale/comments, and an individual report was sent to each expert describing their ratings from the previous round. After rating outcomes in each section (i.e., CCS or other independent prescribing), experts were invited to provide further comments in free-text boxes to share additional reasons for their ratings not reflected in the summarised rationales. A sample of the second-round questionnaire is provided in Appendix 4.

### Third round

In the third round, only outcomes that did not reach consensus after the second round were subject to further rating in the third round. Consensus for inclusion was defined as ≥75% of experts rating an outcome as critically important (scores 7-9), and <25% rating as critically important was the threshold for exclusion. Remaining outcomes were rated again, with second-round summaries presented in the survey and individual reports sent to experts. Outcomes rated by ≥75% of experts as critically important in the third round were deemed to reach consensus, those rated by 25-74% as “Medium-priority,” and <25% as “Low-priority.”

Additionally, an extra set of questions was included, after the rating of important for outcomes for CCS. For each outcome already deemed included or still under consideration in the third round, experts rated the feasibility of measuring each outcome using existing health data sources or systems in Ireland via a 5-point Likert scale (strongly disagree to strongly agree) and suggested potential data sources in a free-text box (with or without any modifications to the source or system). As described above, only these questions were completed by additional experts in health data sources and systems recruited for the third round.

### Patient and Public Involvement

The research team consisted of academics, including those with healthcare professional backgrounds, and PPI contributors, identified from the PPI sub-committee of the Irish Expert Taskforce. Through research team meetings, PPI contributors were involved through all study phases, including refining the study aim and methodology, reviewing and revising study recruitment materials and surveys, and considering changes to outcome wording and description based on Delphi panel comments. Finally as members of the research team, PPI contributors co-authored this paper.

### Statistical analysis and consensus definition

Data analysis and visualisation were conducted using Microsoft Excel (Microsoft Corporation, Redmond, WA), R-Studio software and Python 3. Responses to each outcome were summarised using measures of central tendency (mean, median), spread (25th and 75th percentile) and percentage of experts rating as critically important (i.e. a score of 7 or higher) or not important (i.e. a score of 3 or less). Consensus as critically important was based on ≥75% of experts rating the outcome as critically important. The threshold of 75% is frequently used for Delphi studies, and sets a relatively high threshold for inclusion of outcomes, which is appropriate given the importance of prioritising outcomes to measure and assess impact ^19^. The responses in the free-text comments were analysed and aggregated across all expert groups by two independent reviewers (AHA and FM).

## Results

### Participants

A total of 35 experts responded to the first round of the study (response rate 83%), including five members of the public and patients. Of them, 33 completed the second round and 30 completed third. A further three new experts in health data sources/systems (of five invited, response rate 60%) also completed the third-round questions on feasibility of outcome measurement (see Figure 1).

**Figure 1.**
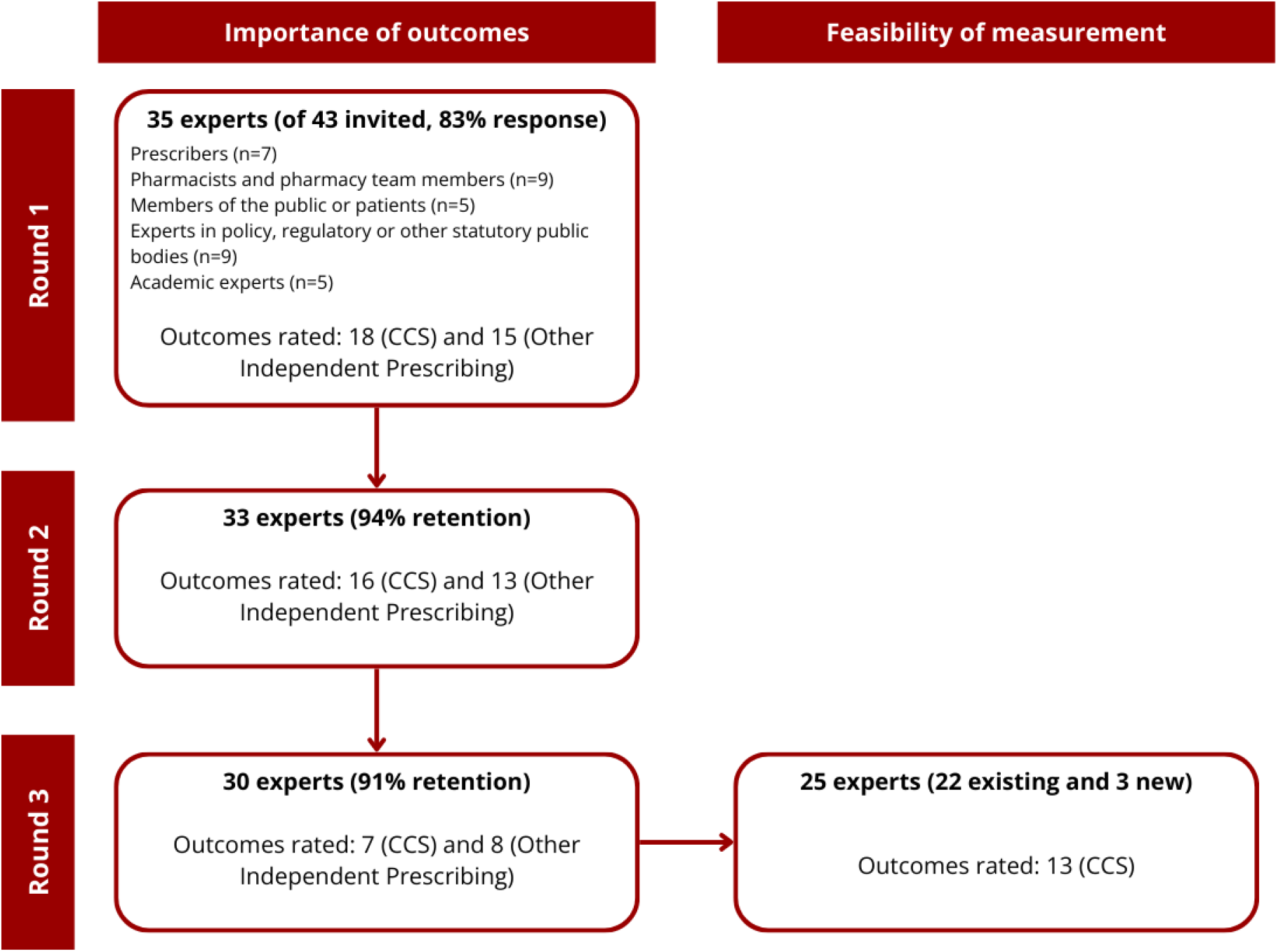
Flow diagram of experts participating in the Delphi study *CCS: Common Condition Service*

### Importance of outcomes

After the second round, six of 18 outcomes for pharmacist prescribing within a CCS achieved consensus as critically important (75% or more rating as critically important), and one further outcome after the third round. These were symptom resolution or improvement/clinical cure, referral to other healthcare providers, re-consultation with other health care providers/other healthcare utilisation, guideline concordance/appropriateness of medications, patient experience and satisfaction with care, cost of care to the healthcare system, and access to care (see Table 1 and Figures 2-3).

**Figure 2.**
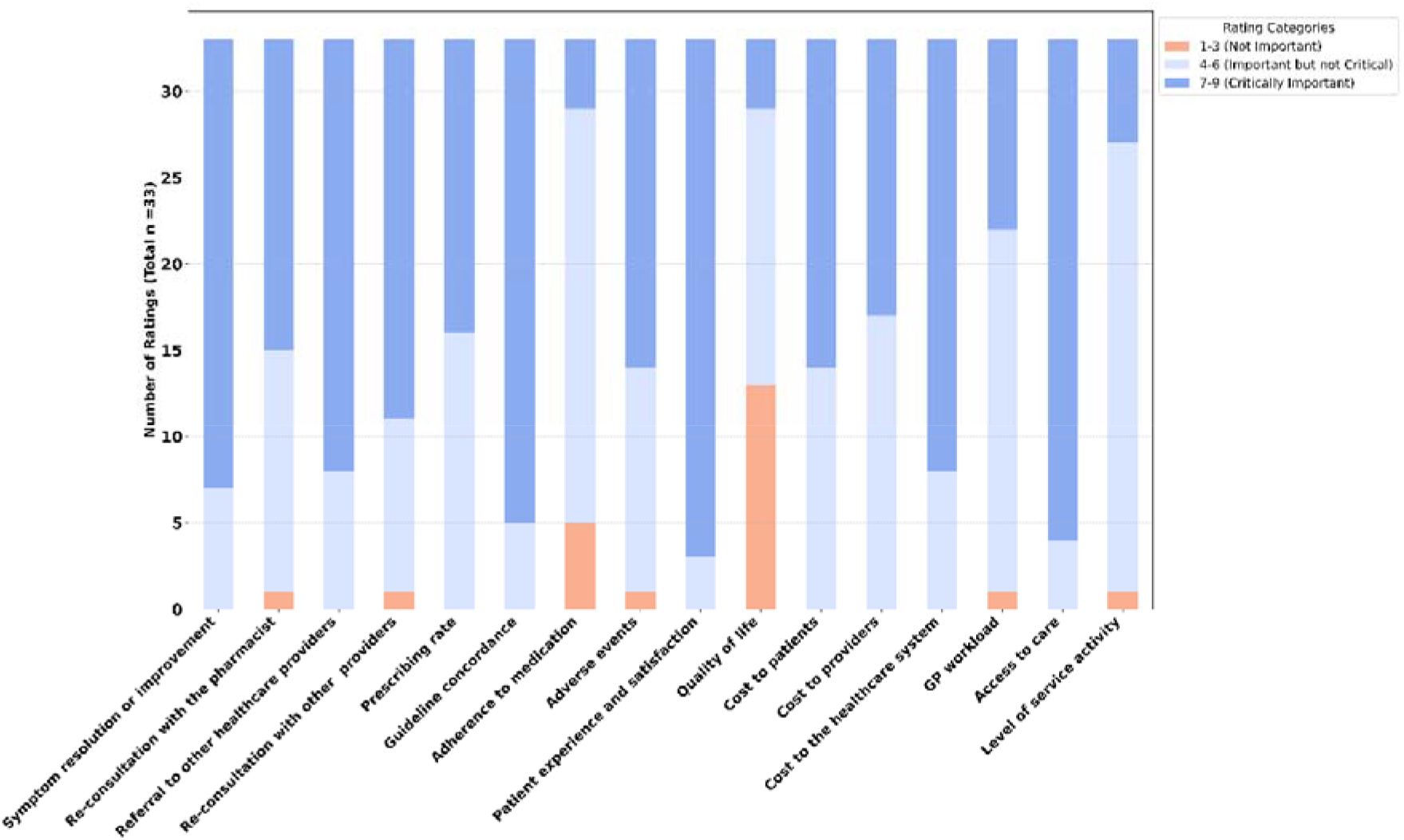
Distribution of second-round ratings for outcomes evaluating the impact of pharmacist prescribing within a Common Conditions Service

**Figure 3.**
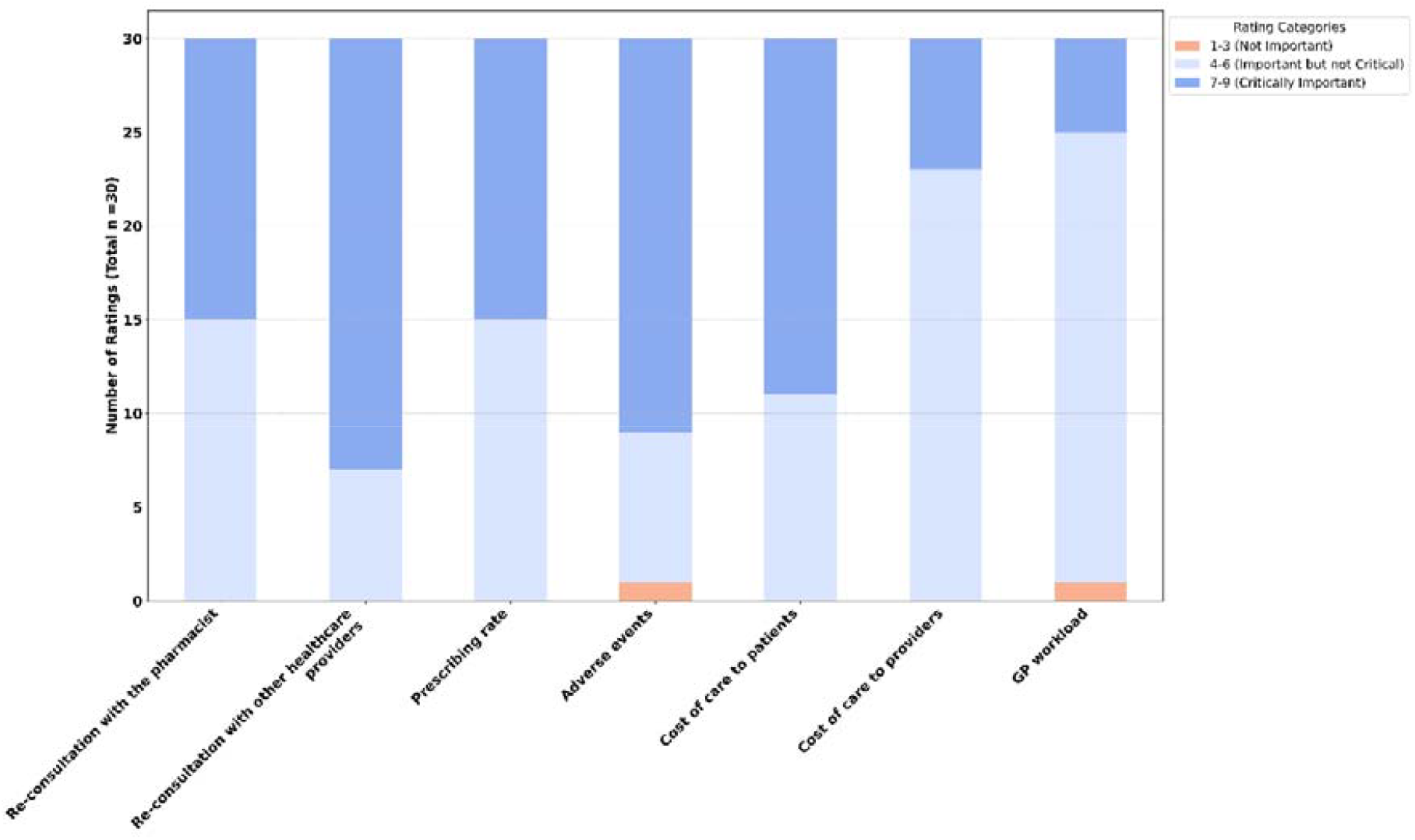
Distribution of third-round ratings for outcomes evaluating the impact of pharmacist prescribing within a Common Conditions Service

For other independent pharmacist prescribing, four of 15 outcomes achieved consensus as critically important after the second round, and a further six outcomes after the third round: clinical effectiveness, guideline concordance/appropriateness of medications, patient experience and satisfaction with care, cost of care to service providers, adverse events, cost of care to the healthcare system, mortality, healthcare utilisation, cost of care to patients, and access to care (see Table 1 and Figures 4-5). Full summaries of the central tendency and distribution of expert rating statistics are provided in Appendices 5–7. Furthermore, Appendix 8 shows how the percentage rating as critically important shifted for each outcome across rounds.

**Figure 4.**
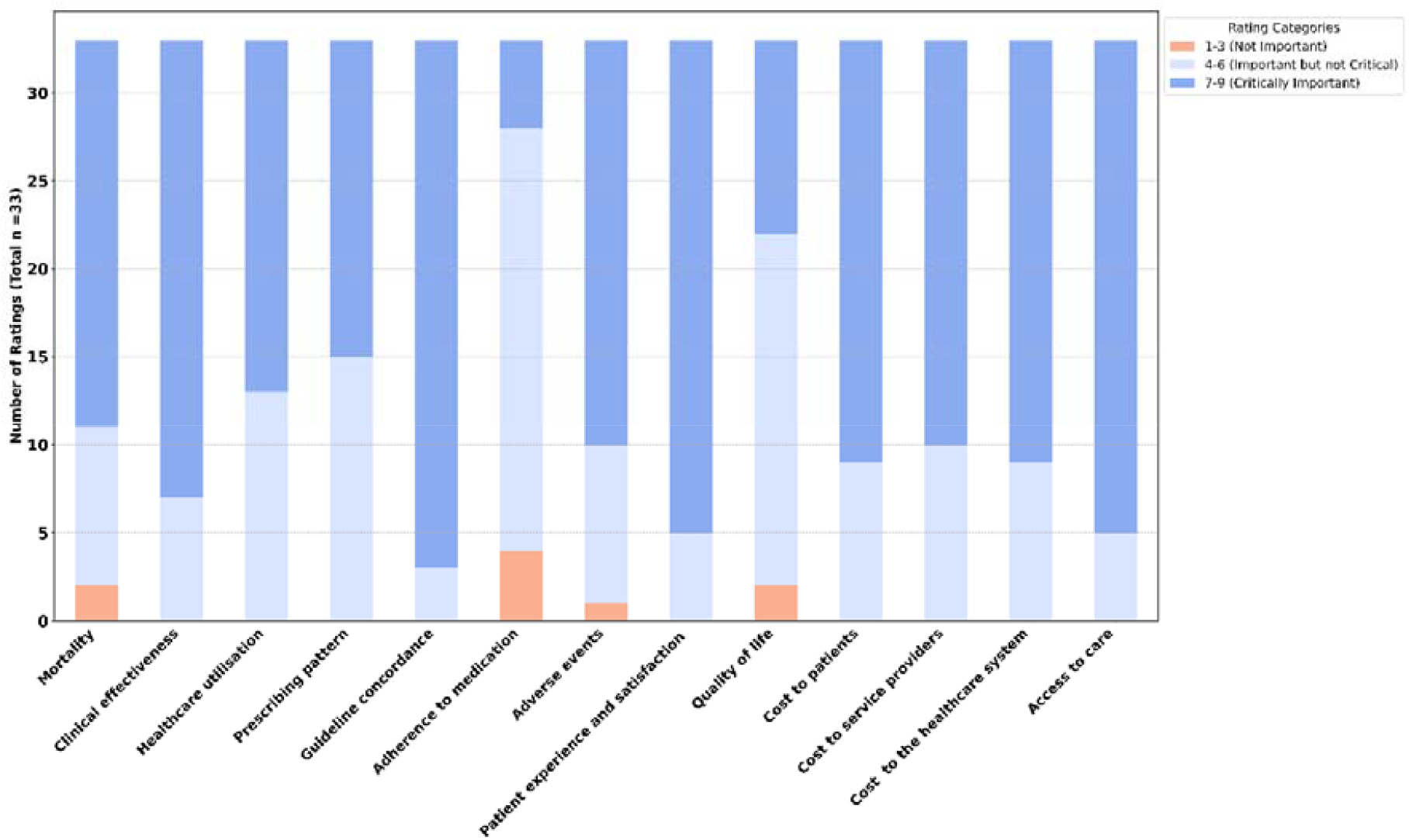
Distribution of second-round ratings for outcomes evaluating the impact of other independent prescribing

**Figure 5.**
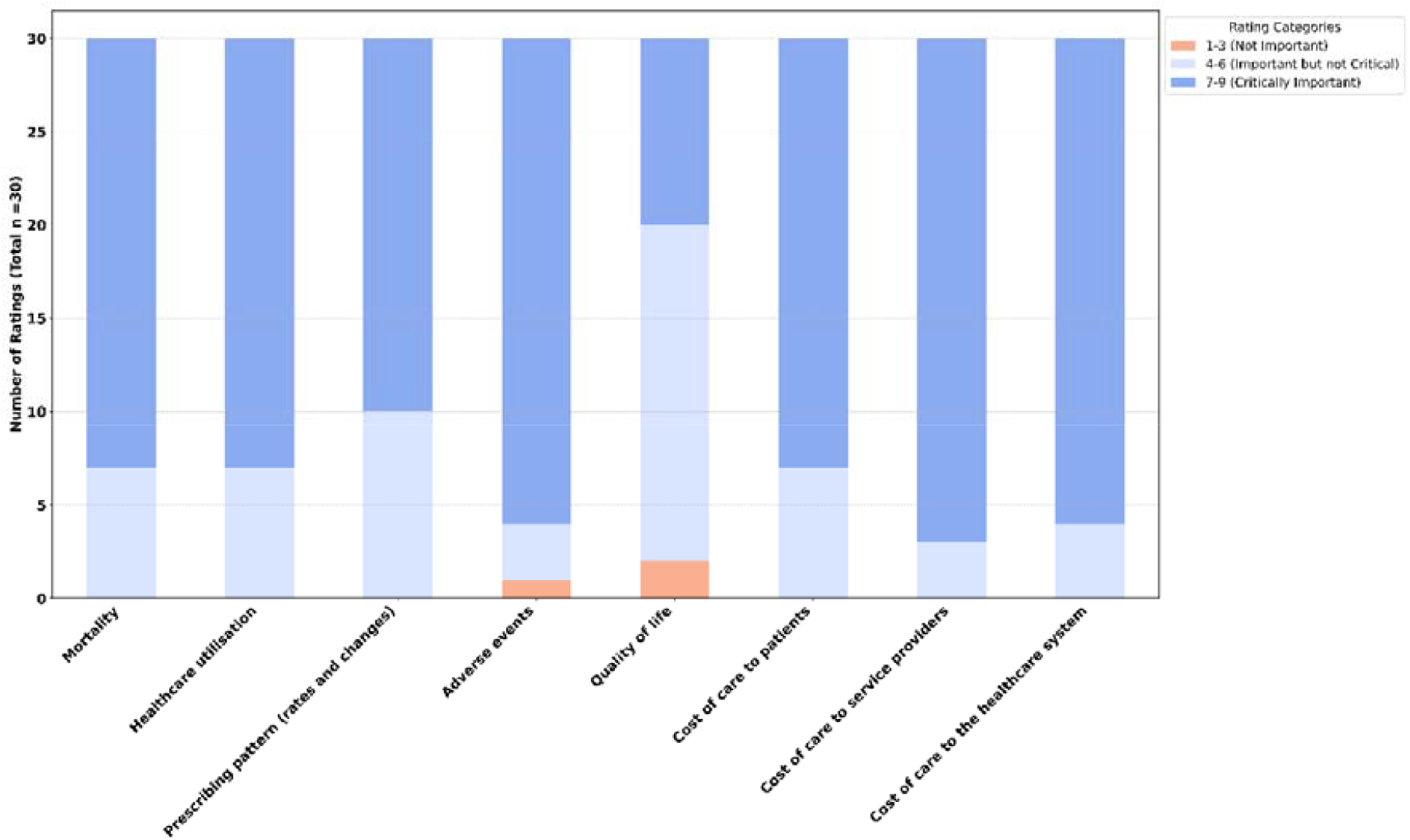
Distribution of third-round ratings for outcomes evaluating the impact of other independent prescribing

Comments from the expert panel, including members of the public and patient contributors, highlighted that the outcomes rated as important have the potential to serve as key audit measures or performance indicators. These outcomes were perceived as instrumental in promoting evidence-based practice, enhancing patient safety, supporting future pharmacist development and learning, and to demonstrate adherence to the scope of practice and awareness of limitations, which are important aspects of the governance of good prescribing.

Comments against the importance of outcomes often focused on the practicality of measuring and interpreting them (e.g. due to lack of an appropriate comparator or denominator). Some outcomes were seen as complex, influenced by numerous factors beyond the direct control of pharmacist prescribing. Therefore, some experts felt these outcomes were hard to interpret and may not reliably reflect the quality or appropriateness of care. The summary of comments from the first two rounds regarding the importance of outcomes is presented in Appendix 9, with samples of representative quotations.

### Feasibility of measuring important outcomes in a CCS

Figure 6 shows the distribution of 25 experts’ responses regarding the feasibility of collecting or assessing outcomes for evaluating pharmacist prescribing within a CCS using existing health data sources or systems in Ireland (with or without modification). “Prescribing rate” (76%) and “Cost of care to patients” (64% had the highest agreement (i.e. strongly agree or agree) for the feasibility of their measurement. Less agreement (around 50% of expert panel) was reported for outcomes such as “Cost of care to providers”, “Cost of care to the healthcare system”, “Re-consultation with the pharmacist”, “Referral to other healthcare providers”, and “Guideline concordance”. In contrast, “Re-consultation with other healthcare providers” and “Symptom improvement or clinical cure” had the lowest feasibility ratings.

**Figure 6.**
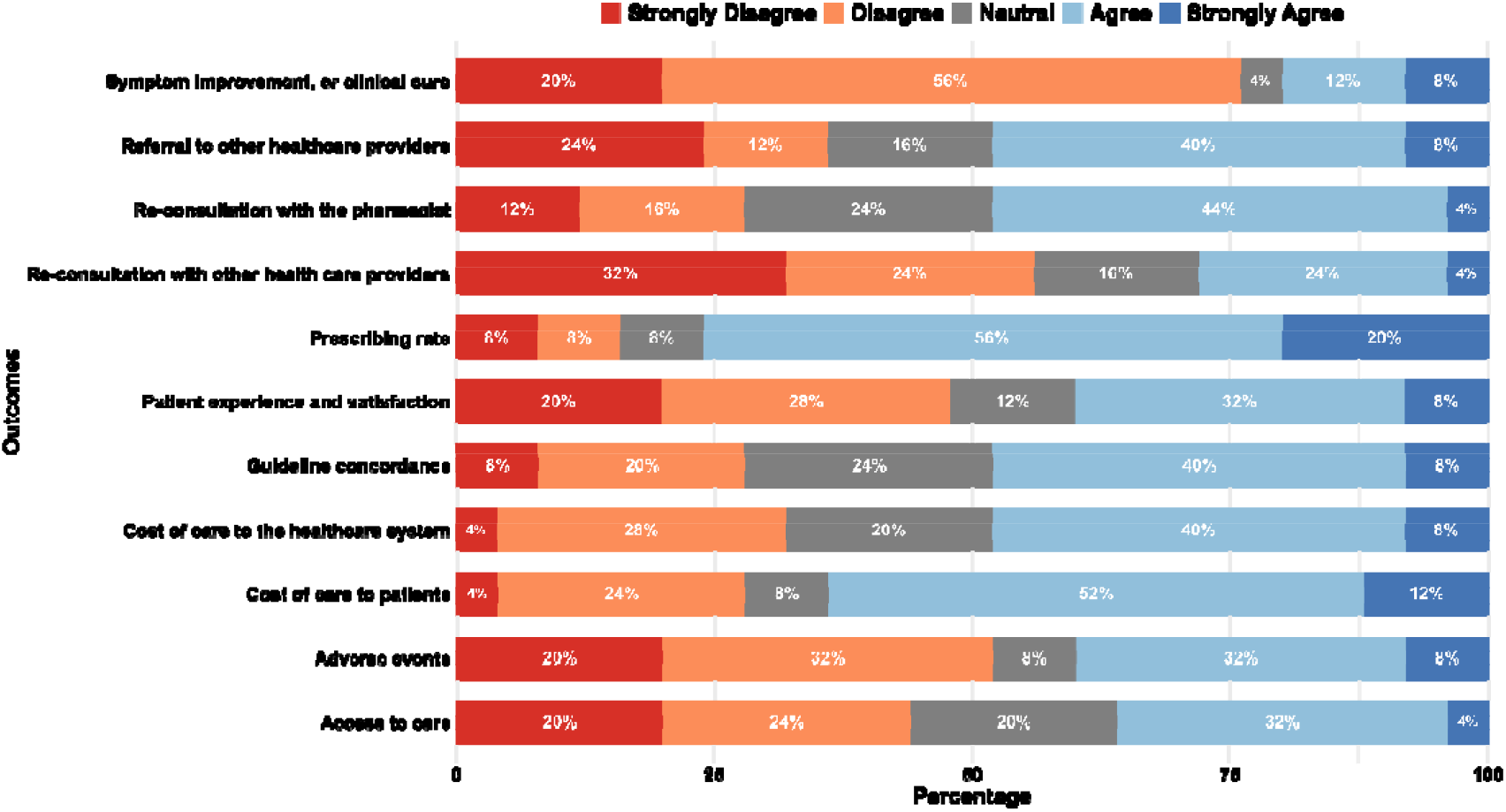
Level of agreement with feasibility of measuring outcomes using existing health data sources or systems in Ireland.

In comments, the experts identified significant limitations in current pharmacy software for measuring pharmacist prescribing outcomes in CCSs in Ireland. Experts emphasised that no standardised approach exists, necessitating interim solutions while longer-term systems are developed. They suggested developing a new digital solution specific to a CCS; or adapting the current IT infrastructure and care pathways to capture required data (e.g. adoption of standardised clinical coding systems such as SNOMED and the integration of unique patient identifiers). Experts recommended measuring outcomes using existing pharmacy systems (e.g. PMR), and other external data sources/set such as national healthcare reimbursement records and adverse reaction reporting databases. For measuring outcomes that need prospective data collection, experts suggested direct patient data collection methods via digital surveys, follow-up contact (e.g. phone calls) or embedded tools in pharmacy systems (e.g. patient portals or patient-friendly interfaces). Appendix 10 summarises in more detail the expert panel’s views on how to measure important outcomes, as explored in the additional questions in the third round. Table 2 outlines critically important and medium-priority outcomes, and potential data sources for common conditions after concluding the three rounds of the Delphi study.

**Table 2.** Critically important outcomes and medium-priority outcomes for evaluating the impact of pharmacist prescribing within a Common Conditions Service.

| Outcomes | % rating<br>as | %<br>Agreement | Potential existing health data sources/<br>systems identified by experts |
| --- | --- | --- | --- |

|  | critically<br>important | with<br>feasibility to<br>measure<br>using<br>existing<br>health data<br>sources or<br>systems | Pharmacy<br>systems | External data<br>sources/set | Direct patient<br>data collection |
| --- | --- | --- | --- | --- | --- |
| **Critically Important** |  |  |  |  |  |
| Patient experience and satisfaction | 90.9 | 40 |  |  | ? |
| Access to care | 87.9 | 36 | ? | ? | ? |
| Guideline concordance/<br>appropriateness of medications | 84.8 | 48 | ? |  |  |
| Symptom improvement, or clinical cure | 78.8 | 20 | ? |  | ? |
| Re-consultation with other health care providers/other healthcare utilisation | 76.7 | 28 | ? | ? |  |
| Cost of care to the healthcare system | 75.8 | 48 |  | ? |  |
| Referral to other healthcare providers | 75.8 | 48 | ? | ? | ? |
| **Medium Priority** |  |  |  |  |  |
| Adverse events | 70.0 | 40 | ? | ? |  |
| Cost of care to patients | 57.6 | 64 | ? | ? | ? |
| Prescribing rate | 50.0 | 76 | ? | ? |  |
| Re-consultation with the pharmacist | 50.0 | 48 | ? | ? |  |

## Discussion

### Summary of main findings

This Delphi study identified critically important outcomes that should be prioritised when evaluating pharmacist prescribing in the provision of CCS and other independent prescribing services. Over three rounds, an expert panel reached consensus on seven outcomes for CCS, with highest ratings for “Patient experience and satisfaction”, “Access to care”, and “Guideline concordance/appropriateness of medications”. Panel experts considered that these outcomes may be amenable to measurement to varying degrees using existing health data sources in Ireland. For other independent prescribing, ten outcomes were deemed critically important (e.g. “Guideline concordance/appropriateness of medications”, “Cost of care to service providers”, and “Adverse events”).

The current findings both align with and extend beyond previous research aimed at evaluating pharmacist prescribing. Our results demonstrate strong agreement with Newham et al.’s nominal group technique study employed to identify key outcomes for pharmacy-led clinical services including prescribing in primary care in Scotland.^16^ Similar to their identification of five key outcomes spanning clinical, economic, humanistic, and service domains, the current study proposed multidimensional evaluation priorities. The prioritisation of patient experience in our study parallels their identification of patient experience as a key humanistic outcome, reinforcing the shift toward patient-centred evaluation metrics – contrasting with earlier literature, which placed greater emphasis on disease indicators (e.g. clinical effectiveness measures such as reduced blood pressure).^16, 20^ However, the Delphi expert panel in the current study deemed clinical outcomes such as symptom resolution and clinical effectiveness as critically important in evaluating pharmacist prescribing in CCSs and other independent prescribing contexts, respectively. The current results are consistent with findings from reviews of the views of stakeholders and experiences of patients and the public, which frequently report on patients’ experience/satisfaction in both minor ailments and other independent prescribing.^15, 21^ Importantly, while patient satisfaction is often used to evaluate service experience and quality for community pharmacy services, growing evidence cautions against over-reliance on such measures.^22^ By also prioritising cost and access-related measures alongside other patient-reported outcomes (e.g. effectiveness, adverse events), the current study contributes to this growing emphasis on more comprehensive, person-centred approaches in assessing service users’ experience and satisfaction.

There was a differential importance of safety metrics between contexts, with “adverse events” achieving critically important status in other independent prescribing but only medium priority for CCS. This represents a novel insight extending beyond the findings of Newham et al. which emphasised medication-related adverse events as a primary clinical prescribing related-outcome but did not identify context-specific variations in the importance. While Newham et al. identified medication optimisation as a core service outcome, our findings similarly emphasised guideline concordance as critically important across both settings (84.8-90.9% rated as critically important). The strong emphasis on the importance of guideline concordance/appropriateness of medications across both settings validates the findings from several reviews that identify adherence to clinical guidelines as a frequently used outcome to evaluate pharmacist prescribing.^23, 24^

Identification of “cost of care to the healthcare system” as critically important in both contexts and “cost of care to patients” as critically important in other independent prescribing context and of medium priority in CCSs aligns with findings in existing literature, for example Newham et al.’s inclusion of cost-effectiveness among their five key outcomes. Moreover, Jebara et al. (2018) review of stakeholders’ views of pharmacist prescribing highlighted that pharmacists and physicians positively perceived this expanded professional role across postimplementation studies with benefits including better and continuous patient care and management and reduced cost of therapy.^15^ However, our finding that “cost of care to providers” had differential consensus on importance across context (low priority in CCs, critically important for other independent prescribing) represents a novel contribution not specifically captured in other studies. This may be due to community pharmacies, who are independent businesses and contractors, representing the providers in the former case, with the latter case also including hospitals, general practices, and other providers potentially. In both cases, this cost-related outcome remains a relevant consideration, but the specific concerns and perceived impact of the service may differ depending on how pharmacist prescribing services are delivered and funded in each setting. The critical importance of “access to care” in this Delphi study contributes to the broader body of evidence suggesting that improving access to care is a central objective of implementing pharmacist prescribing, in line with conclusions drawn from multiple reviews and studies incorporating diverse stakeholder perspectives.^25, 26^

### Outcome importance depends on context of pharmacist prescribing

While Newham et al.’s considered health-related quality of life as one the five key outcomes, experts in the current Delphi frequently considered outcomes less important if there were concerns about valid interpretation and where there were multiple, strong factors that may influence the outcome beyond pharmacists’ prescribing. This varied by context, for example with quality of life being rated higher for other independent prescribing compared to prescribing within a CCS, and experts’ comments suggested the typically short timeframe of common conditions would give little opportunity to impact on quality of life. This may have contributed to the higher number of outcomes reaching the threshold for critical importance for other independent prescribing. Furthermore, the broader scope and different settings, may also have contributed to the higher number of critically important outcomes in this context.

The other contextual factor that was highlighted repeatedly in expert comments was entitlement to and reimbursement for pharmacist prescribing within a CCS for individuals covered by public health schemes. This was noted as a factor that would affect the importance of some outcomes (i.e. that cost outcomes would have greater importance if the service was publicly funded), and also impact on the ease of measurement (i.e. whether reimbursement claims would be a source to capture relevant data). Most importantly, entitlement and reimbursement were noted to potentially influence the impact of pharmacist prescribing on access to care, with experts emphasising that a service only available to individuals with the ability to pay would undermine equity of access and reinforce existing disparities in timely care.

### Importance of assessing outcomes in a robust way

The findings of this research emphasise the importance of data infrastructure, both harnessing and enhancing existing systems, and ensuring that future systems are interoperable, adhere to relevant standards, use consistent and comprehensive coding, and incorporate unique patient identifiers. Experts emphasised the importance of ensuring data captured within such infrastructures is shareable and useable for the purposes of evaluation, research and quality improvement to ultimately improve healthcare delivery and outcomes. This is particularly given developments such as the European Health Data Space Regulation.^27, 28, 29, 30^

.Experts strongly emphasised that the impact of pharmacist prescribing can only be validly assessed using routine data with appropriate design and analysis of evaluations. This requires an appropriate comparison, for example against other prescribing healthcare professionals or baseline data prior to the introduction of pharmacist prescribing. In some cases, critical outcomes identified in this research may not be currently measured for prescribing by other healthcare professionals or before introducing pharmacist prescribing. While this should not preclude measuring outcomes like these for other purposes (e.g. describing delivery of service, or providing feedback to pharmacists to support confidence and development), resources and efforts may be better focused on areas where robust comparisons and interpretation of outcomes is possible to assess the true impact of pharmacist prescribing.

### Strengths and limitations of the methodology

A significant strength of this study is its methodological distinction between prescribing contexts, which enables a more nuanced assessment of outcome relevance in accordance with the complexity and scope of prescribing. The Delphi methodology is appropriate for reaching consensus among a diverse group of experts and limiting potential for hierarchy to influence individual judgements. There was strong representation across different interest-holder groups and retention across rounds (>90% participating at subsequent rounds), and for each outcome, most experts provided comments alongside their ratings. In particular, members of the public and patients were included in the Delphi panel, ensuring that the patient voice was captured as end-users of prescribing services. Furthermore, members of PPI sub-committee of the Expert Taskforce were part of the research team, contributing to the study design, refinement of outcome lists, and interpretation of findings, further ensuring the relevance of the study and its findings to patients and the public.

The questions relating to measurement were not subject to repeated rounds of rating, and so the agreement for these responses represents the views of the experts without the opportunity to reach group consensus. The feasibility of measurement of outcomes using existing health data sources was not assessed for other independent pharmacist prescribing, due to the heterogeneity of contexts/settings this may involve. However, recommendations on measurement relating to the CCS may apply to other independent prescribing in the community pharmacy setting. This approach may have reduced burden to participants and mitigated attrition, as Delphi studies with a higher number of items may be associated with significantly lower response rates in subsequent rounds.^31^ Additionally, the ratings of feasibility of measurement were based on expert judgement and may not reliably reflect the actual feasibility of measuring certain outcomes. These ratings may have been affected by experts’ perceptions of the accessibility of relevant data within existing health sources and systems. However, this provides a basis for future work engaging software vendors and dataholders to objectively determine the feasibility of extracting necessary data to measure these outcomes. While the study was conducted in the Irish setting, the outcome importance is unlikely to be significantly influenced by local considerations and so may be generalisable to other contexts.

## Conclusions

Seven outcomes reached consensus as critically important for assessing the impact of pharmacist prescribing within a CCS in community pharmacies, with ten outcomes for other independent pharmacist prescribing contexts. These covered clinical, safety, economic, and patient-reported outcomes. While some of these outcomes can be measured using existing health data sources in Ireland, substantial modifications are necessary to improve the comprehensiveness and interoperability of these systems. These findings can provide guidance for prioritising outcome measurement in routine or specific evaluations of pharmacist prescribing in other jurisdictions.

## Data Availability

All relevant study data are available in the manuscript and supplementary files

## Acknowledgments

The authors gratefully acknowledge the time provided by all those experts who participated in the Delphi study.

## Notes

**Declaration of interests** JS was a Member of the Expert Taskforce to support the expansion of the role of pharmacists in Ireland. CMC was a Member of the Research Sub-Committee of the Expert Taskforce to support the expansion of the role of pharmacists in Ireland. Other authors have no interests to declare.

### Competing Interest Statement

JS was a Member of the Expert Taskforce to support the expansion of the role of pharmacists in Ireland. CMC was a Member of the Research Sub-Committee of the Expert Taskforce to support the expansion of the role of pharmacists in Ireland. Other authors have no interests to declare

### Author Declarations

Ethical approval was granted by the RCSI University of Medicine and Health Sciences Research Ethics Committee (REC) (reference number REC202411038)

