## Supplementary Materials for "Identifying and measuring important outcomes for evaluating the impact of pharmacist prescribing in Ireland: A modified Delphi study"

### **Appendix 1: Development of list of outcomes**

The rapid overview of reviews identified a total of 14 outcomes for evaluating pharmacist prescribing in minor ailments and common conditions, and 12 outcomes were identified for other forms of pharmacist prescribing. These outcomes were categorised into four domains: clinical outcomes, drug-related and prescribing outcomes, patient-reported and experience outcomes, and economic and other related outcomes. These outcomes were assessed for suitability by the research team (which comprised those with clinical/academic experience of pharmacy, general practice, and evidence synthesis, and patient and public involvement contributors). To operationalise these for rating within the Delphi, modifications were made as follows:

1. Integrate “treatment failure” into “symptom resolution or improvement, or clinical cure”, as the former was regarded as the absence of the latter.
2. Divide re-consultation with the pharmacist and referral to other healthcare providers into separate outcomes as these were regarded as distinct.
3. Divide the quality of life related outcomes into quality of life, general health status, and patient functioning, as these were regarded as distinct.
4. Divide the “cost of service delivery” outcome into three outcomes representing costs to the patient, provider of the service, and healthcare system, as it was regarded that importance of costs may differ based on who bears the costs.
5. Remove the “resource use due to healthcare utilisation”, as this was regarded to overlap fully with the “healthcare utilisation” and cost-related outcomes.

These modifications yielded 18 outcomes for evaluating pharmacist prescribing within common condition services and 15 for other independent prescribing contexts, which were rated for their importance in the first Delphi round.

### **Appendix 2: Outcomes and descriptions presented to experts, incorporating changes based on expert panel comments**

| **Outcome** | **Description** |
| --- | --- |
| **Common Conditions Service** | |
| Symptom resolution or improvement, or clinical cure | Resolution of the condition/symptoms, or positive changes in symptoms from baseline in terms of severity, frequency and/or duration. |
| Re-consultation with the pharmacist | Returning to the pharmacist for the same condition, related symptoms, or adverse effects after the initial consultation. |
| Referral to other healthcare providers | The pharmacist refers the patient back to another healthcare provider for further evaluation or treatment. |
| Re-consultation with other health care providers/other healthcare utilisation | Seeking care from other healthcare providers or utilising other healthcare services (e.g. emergency department visits or hospitalisations) after the initial consultation or follow-up visits with the pharmacist relating to the same condition/symptoms due to, for example, treatment failure, unresolved symptoms, or prescribing-related issues. |
| Prescribing rate | Prescribing rates at level of individual pharmacists or all pharmacists relative to overall prescribing rates of specific medicines or drug classes (e.g. antimicrobial agent for urinary tract infection (UTI), corticosteroid nasal spray for allergic rhinitis). |
| Guideline concordance/appropriateness of medications | Pharmacists' treatment following clinical guidelines or protocols, and the appropriateness of prescribing (e.g. appropriate drug selection, dose and frequency), acknowledging the role of individual patient’s goal of therapy, preferences, etc. |
| Patient adherence to medication | How well a patient’s behaviour aligns with the prescribed instructions for taking medication to achieve therapeutic objectives. |
| Adverse events | Adverse clinical outcomes associated with pharmacist prescribed medications (e.g. drug related adverse effects such as stomach upset) or the consultation (e.g. medications allergy after not confirming allergies). |
| Patient experience and satisfaction with care | Experience and satisfaction overall, or with aspects such as access to care (e.g. easiness and convenience), interpersonal communication (e.g. shared decision making), continuity and coordination, comprehensiveness of services, and trust in healthcare providers. |
| Quality of life* | Perceived quality of a person’s daily life, assessing their well-being or lack thereof. This includes all emotional, social, cognitive and physical aspects of the individual's life, and may focus on how a person’s well-being and perceived general health is impacted by a condition or disease, and their ability to perform usual activities (incorporating general health status and functioning since the first round). |
| Cost of care to patients | Direct and indirect costs to patients of accessing pharmacist prescribing, on its own, relative to alternatives, or relative to the time to access care. |
| Cost of care to providers | Cost-effectiveness and cost minimisation (considering direct and indirect costs) from the perspective of community pharmacies providing the common condition service. |
| Cost of care to the healthcare system | Cost-effectiveness and cost minimisation (considering direct and indirect costs) from the perspective of the healthcare system. |
| GP workload | Increase or decrease in number or length of consultations with general practice staff |
| Access to care | Measures such as the proportion of eligible people having a consultation or receiving medicines, the number of overall dispensed prescriptions, and time to receipt of prescriptions, equity of access and perceived convenience of accessing care. |
| Level of service activity | Overall volume of patients availing of the service, number of prescriptions issues, or the numbers of service claims. |
| **Other Independent Prescribing** | |
| Mortality | Death from any cause or specific causes. |
| Clinical effectiveness | Measures of the specific benefits of prescribed treatments, such as improvement/control of blood pressure, blood glucose and blood cholesterol, reduction in major cardiovascular event risk, prevention of unplanned pregnancies, HIV prophylaxis, etc |
| Healthcare utilisation | Increased or decreased primary care visits, emergency department visits, or hospitalisations. |
| Prescribing pattern (rates and changes) | Rates of prescribing of specific medicines or drug classes, changes made to prescribed treatment, or initiation or discontinuation of medicines. |
| Guideline concordance/appropriateness of medications | Pharmacists’ treatment following clinical guidelines or protocols, and the appropriateness of prescribing (e.g. appropriate drug selection, dose and frequency, prescribing omitted medications, discontinuing inappropriate medications). |
| Patient adherence to medication | How well a patient’s behaviour aligns with the prescribed instructions for taking medication to achieve therapeutic objectives. |
| Adverse events | Adverse clinical outcomes associated with both pharmacist-prescribed medications (e.g. hypertension from rapid antihypertensives titration) or the intervention (e.g. chest pain overlooked/misdiagnosed as heartburn). |
| Patient experience and satisfaction with care | Experience and satisfaction overall, or with aspects such as access to care (e.g. easiness and convenience), interpersonal communication (e.g. shared decision making), continuity and coordination, comprehensiveness of services, and trust in healthcare providers. |
| Quality of life* | Perceived quality of a person’s daily life, assessing their well-being or lack thereof. This includes all emotional, social, cognitive and physical aspects of the individual's life, and may focus on how a person’s well-being and perceived general health is impacted by a condition or disease, and their ability to perform usual activities (incorporating general health status and functioning since the first round). |
| Cost of care to patients | Direct and indirect costs to patients of accessing pharmacist prescribing, on its own, relative to alternatives, or relative to the time to access care. |
| Cost of care to service providers | Cost-effectiveness and cost minimisation (considering direct and indirect costs) from the perspective of institutions providing the care (e.g. community pharmacies, GP practices, hospitals). |
| Cost of care to the healthcare system | Cost-effectiveness and cost minimisation (considering direct and indirect costs) from the perspective of the healthcare system. |
| Access to care | Measures such as the proportion of eligible people receiving medicines, the number of overall dispensed prescriptions, time to receipt of prescriptions, equity of access and perceived convenience of accessing care. |

* Quality of life incorporates general health status and patient functioning, as revised from the second round onwards.

### **Appendix 3: Sample of the first round survey**

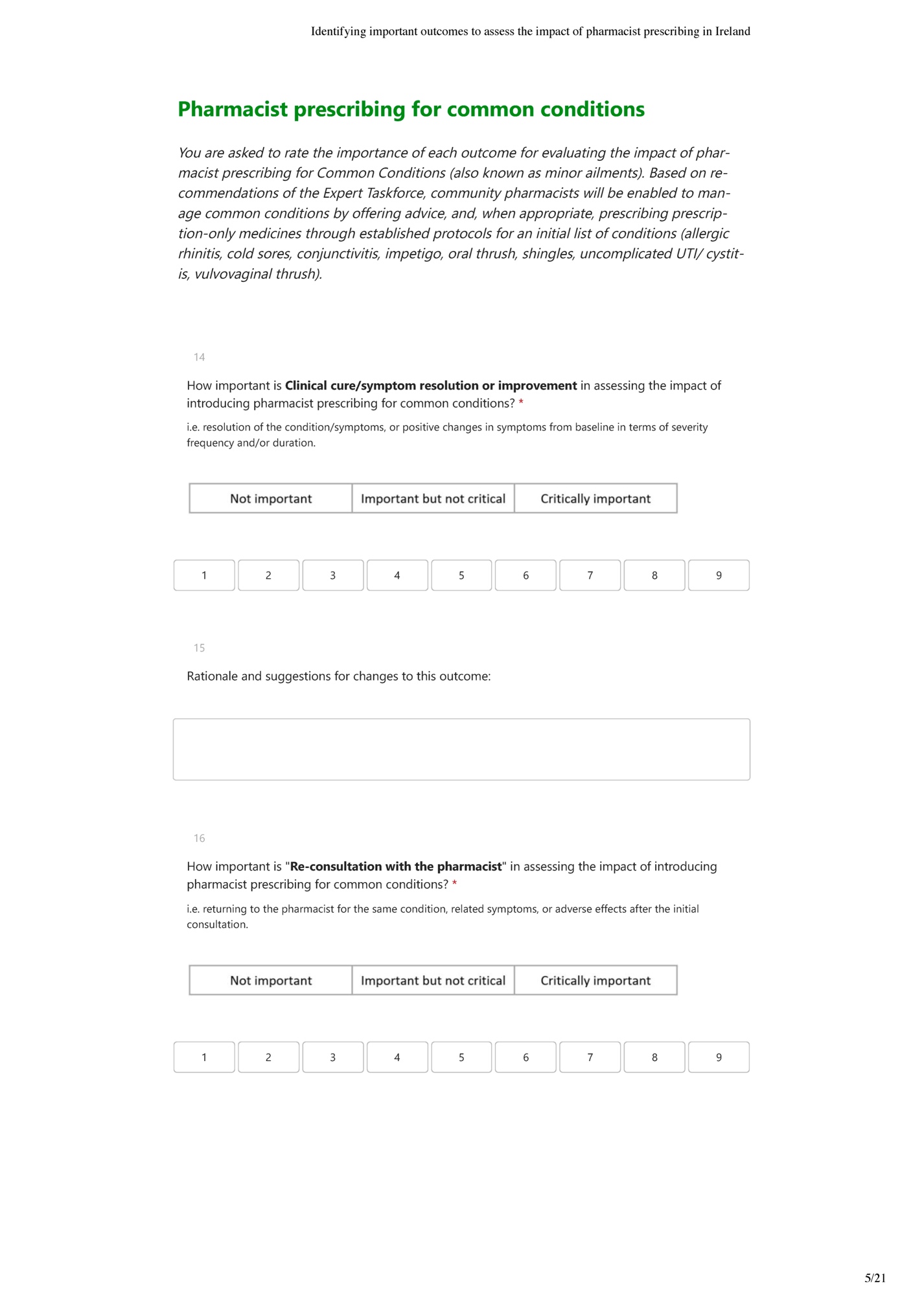

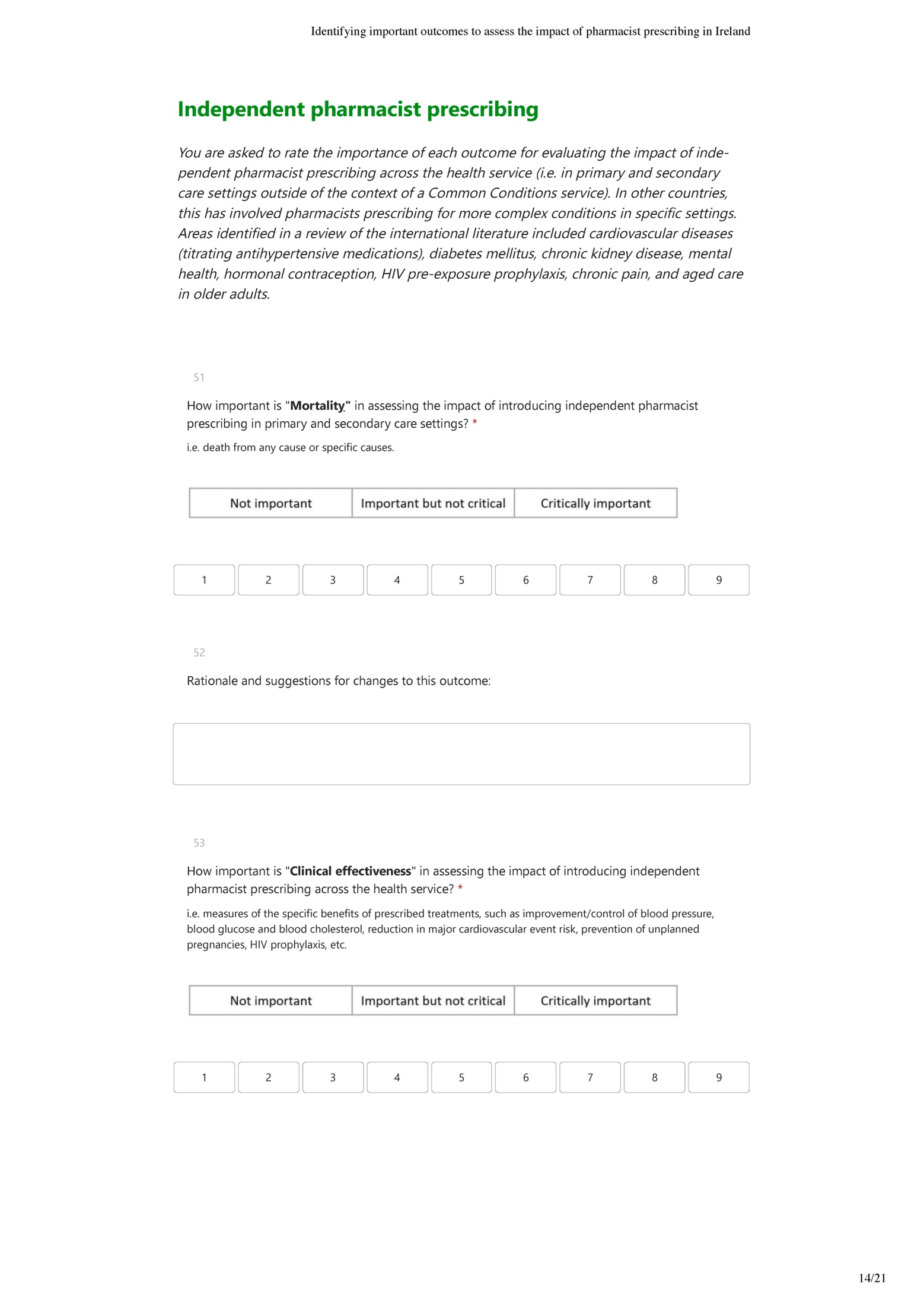

### **Appendix 4: Sample of the second round survey**

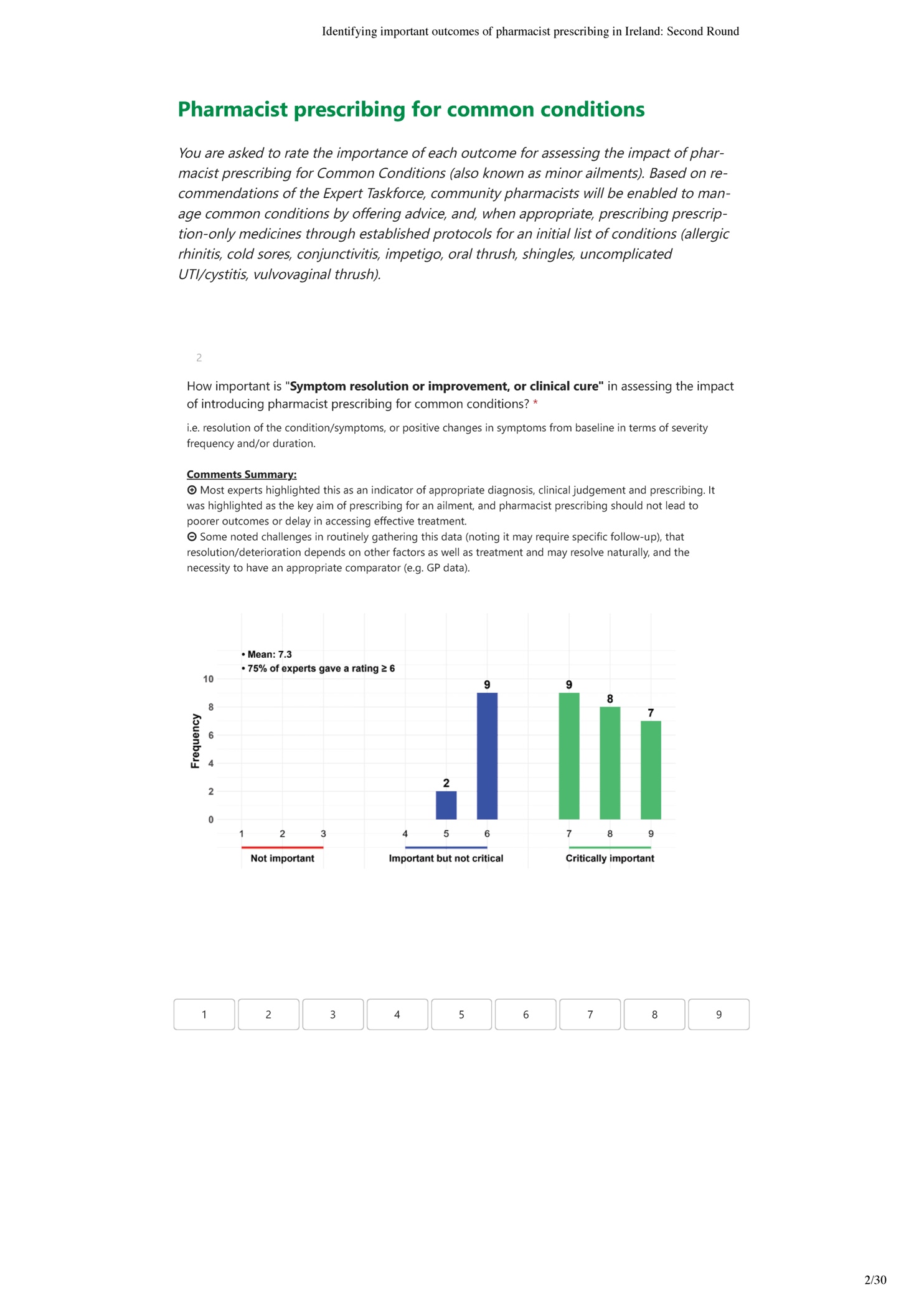

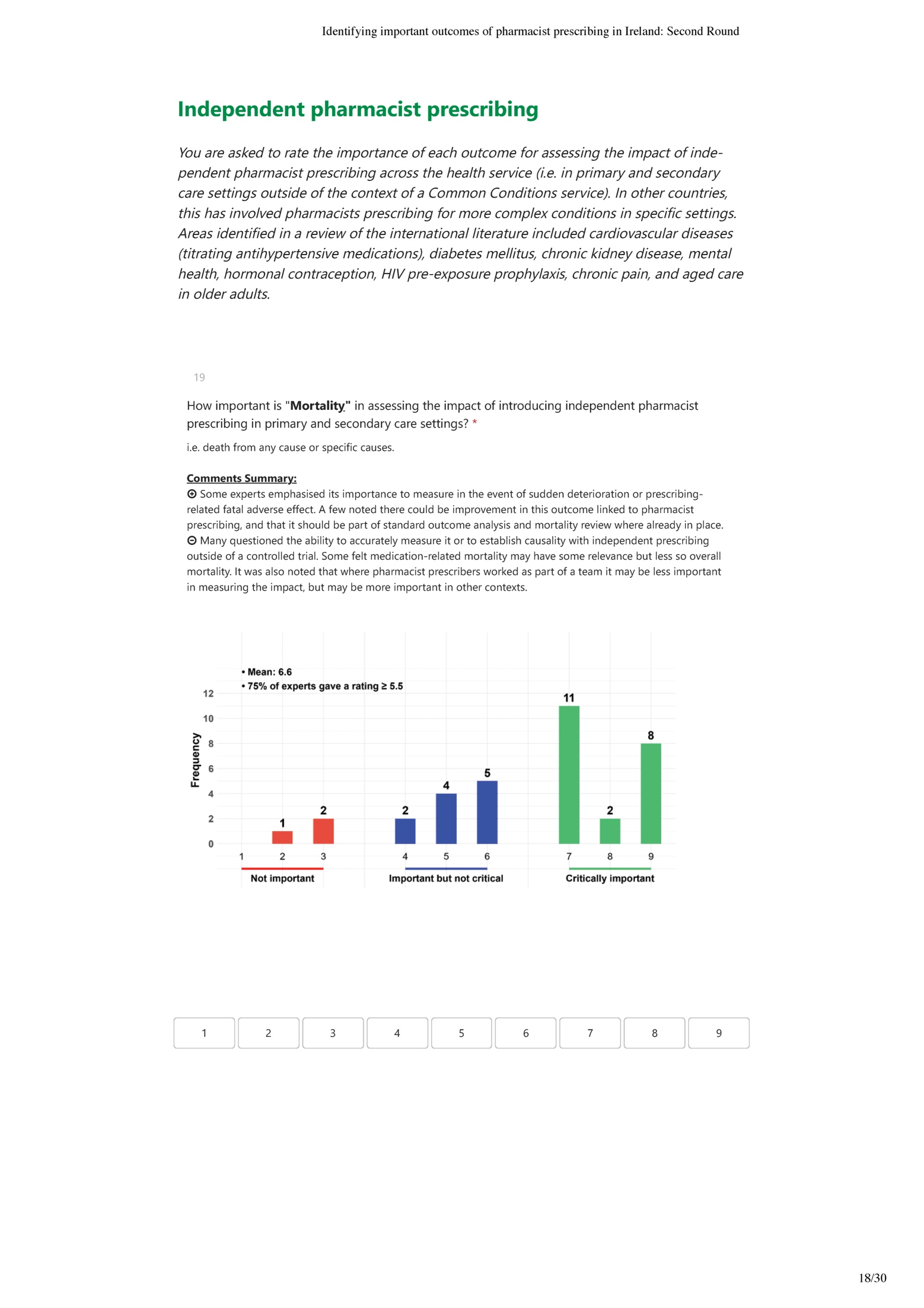

### **Appendix 5 Results of the first round (importance summaries)**

18 outcomes were subject to rating in the first round for evaluating pharmacist prescribing within a Common Conditions Service. Two of them (i.e. patient experience, and guideline concordance) were rated as critically important by more than 75% of the expert panel. Quality of life and the related outcomes received the lowest ratings, with less than 25% of the expert panel rating them as critically important). For other independent prescribing, 15 outcomes were included in the first round, and one outcome (i.e. guideline concordance) out of 15 outcomes was rated as critically important by more than 75% of the expert panel.

Based on experts’ feedback, several outcome titles and descriptions were revised for the second-round survey (see Table 1 footnote). “Symptom resolution” was reworded to reduce emphasis on clinical cure, and “Adverse effects” was renamed “Adverse events” to reflect a broader scope. Due to high correlation of ratings and expert feedback, “General health status” and “Patient functioning” were incorporated under the “quality of life” outcome for subsequent rounds,” with only minor adjustments to its description.

| **Outcomes** | **Median** | **Mean** | **25^th^ Percentile** | **75^th^ Percentile** | **% rating of 1-3** | **% rating of 7-9** |
| --- | --- | --- | --- | --- | --- | --- |
| **Common Conditions** | | | | | | |
| Clinical cure/symptom resolution or improvement | 7 | 7.3 | 6 | 8 | 0 | 68.6 |
| Re-consultation with the pharmacist | 7 | 6.4 | 6 | 7 | 5.7 | 54.3 |
| Referral to other healthcare providers | 7 | 7.3 | 6.5 | 8 | 0 | 74.3 |
| Re-consultation with other health care providers/other healthcare utilisation | 7 | 6.8 | 6 | 8 | 5.7 | 62.9 |
| Prescribing rate | 6 | 6.5 | 6 | 7 | 2.9 | 48.6 |
| Guideline concordance/appropriateness of medications | 8 | 7.6 | 7 | 9 | 0 | 82.9 |
| Patient adherence to medication | 5 | 5.4 | 4 | 6.5 | 17.1 | 25.7 |
| Adverse effects | 7 | 6.8 | 6 | 8 | 2.9 | 57.1 |
| Patient experience and satisfaction with care | 8 | 7.6 | 7 | 9 | 0 | 85.7 |
| Quality of life | 5 | 5 | 3 | 6 | 34.3 | 22.9 |
| General health status | 4 | 4.4 | 3 | 6 | 37.1 | 5.7 |
| Patient functioning | 5 | 4.7 | 3 | 6 | 31.4 | 11.4 |
| Cost of care to patients | 7 | 6.9 | 6 | 8 | 0 | 60 |
| Cost of care to providers | 7 | 6.7 | 6 | 8 | 5.7 | 57.1 |
| Cost of care to the healthcare system | 7 | 7.1 | 6 | 8 | 2.9 | 71.4 |
| GP workload | 6 | 6.2 | 6 | 7 | 8.6 | 42.9 |
| Access to care | 7 | 6.9 | 6.5 | 8 | 2.9 | 74.3 |
| Level of service activity | 6 | 6.3 | 6 | 7 | 5.7 | 40 |
| **Independent Prescribing** | | | | | | |
| Mortality | 7 | 6.6 | 5.5 | 8 | 8.6 | 60 |
| Clinical effectiveness | 7 | 7.4 | 6.5 | 9 | 2.9 | 74.3 |
| Healthcare utilisation | 7 | 6.9 | 6 | 8 | 5.7 | 60 |
| Prescribing pattern (rates and changes) | 7 | 6.8 | 6 | 7 | 0 | 51.4 |
| Guideline concordance/appropriateness of medications | 7 | 7.5 | 7 | 9 | 0 | 82.9 |
| Patient adherence to medication | 6 | 5.7 | 4.5 | 6.5 | 17.1 | 25.7 |
| Adverse effects | 7 | 6.8 | 6 | 7.5 | 2.9 | 62.9 |
| Patient experience and satisfaction with care | 7 | 7.4 | 6.5 | 8.5 | 0 | 74.3 |
| Quality of life | 6 | 5.8 | 4.5 | 7 | 8.6 | 37.1 |
| General health status | 5 | 5.1 | 4 | 6 | 20 | 22.9 |
| Patient functioning | 6 | 5.3 | 4 | 6.5 | 17.1 | 25.7 |
| Cost of care to patients | 7 | 6.8 | 6 | 7 | 0 | 68.6 |
| Cost of care to service providers | 7 | 6.9 | 6 | 7.5 | 0 | 60 |
| Cost of care to the healthcare system | 7 | 7.2 | 6 | 8 | 0 | 68.6 |
| Access to care | 7 | 6.9 | 6 | 8 | 2.9 | 68.6 |

### **Appendix 6 Results of the second round (importance summaries)**

After the second round, six out of 16 outcomes for pharmacist prescribing within a Common Conditions Service achieved consensus as critically important (75% or more rating as critically important). These were symptom resolution or improvement/clinical cure, referral to other healthcare providers, guideline concordance/appropriateness of medications, patient experience and satisfaction with care, cost of care to the healthcare system, and access to care. Three outcomes reached consensus for exclusion with less than 25% of the expert panel rating them as critically important: patient adherence to medication, quality of life, and level of service activity. For other independent pharmacist prescribing, four out of 13 outcomes achieved consensus as critically important: clinical effectiveness, guideline concordance/appropriateness of medications, patient experience and satisfaction with care, and access to care. One outcome, patient adherence to medication, reached consensus for exclusion.

| **Outcomes** | **Median** | **Mean** | **25^th^ Percentile** | **75^th^ Percentile** | **% rating of 1-3** | **%rating of 7-9** |
| --- | --- | --- | --- | --- | --- | --- |
| **Common Conditions** | | | | | | |
| Symptom resolution or improvement, or clinical cure | 7 | 7.4 | 7 | 8 | 0 | 78.8 |
| Re-consultation with the pharmacist | 7 | 6.5 | 6 | 7 | 3 | 54.6 |
| Referral to other healthcare providers | 7 | 7.2 | 7 | 8 | 0 | 75.8 |
| Re-consultation with other health care providers/other healthcare utilisation | 7 | 6.6 | 6 | 7 | 3 | 66.7 |
| Prescribing rate | 7 | 6.7 | 6 | 7 | 0 | 51.5 |
| Guideline concordance/appropriateness of medications | 8 | 7.6 | 7 | 9 | 0 | 84.9 |
| Patient adherence to medication | 5 | 5.1 | 4 | 6 | 15.2 | 12.1 |
| Adverse events | 7 | 6.7 | 6 | 7 | 3 | 57.6 |
| Patient experience and satisfaction with care | 8 | 7.7 | 7 | 9 | 0 | 90.9 |
| Quality of life | 4 | 4.6 | 3 | 6 | 39.4 | 12.1 |
| Cost of care to patients | 7 | 6.8 | 6 | 8 | 0 | 57.6 |
| Cost of care to providers | 6 | 6.7 | 6 | 7 | 0 | 48.5 |
| Cost of care to the healthcare system | 7 | 7.2 | 7 | 8 | 0 | 75.8 |
| GP workload | 6 | 6.2 | 6 | 7 | 3 | 33.3 |
| Access to care | 7 | 7.2 | 7 | 8 | 0 | 87.9 |
| Level of service activity | 6 | 6 | 6 | 6 | 3 | 18.2 |
| **Independent Prescribing** | | | | | | |
| Mortality | 7 | 6.6 | 6 | 7 | 6 | 66.7 |
| Clinical effectiveness | 8 | 7.7 | 7 | 9 | 0 | 78.8 |
| Healthcare utilisation | 7 | 6.7 | 6 | 7 | 0 | 60.6 |
| Prescribing pattern (rates and changes) | 7 | 6.7 | 6 | 7 | 0 | 54.6 |
| Guideline concordance/appropriateness of medications | 7 | 7.6 | 7 | 9 | 0 | 90.9 |
| Patient adherence to medication | 6 | 5.5 | 5 | 6 | 12.1 | 15.2 |
| Adverse events | 7 | 6.8 | 6 | 7 | 3 | 69.7 |
| Patient experience and satisfaction with care | 7 | 7.4 | 7 | 8 | 0 | 84.9 |
| Quality of life | 6 | 5.7 | 5 | 7 | 6.1 | 33.3 |
| Cost of care to patients | 7 | 7 | 6 | 8 | 0 | 72.7 |
| Cost of care to service providers | 7 | 6.9 | 6 | 7 | 0 | 69.7 |
| Cost of care to the healthcare system | 7 | 7.2 | 6 | 8 | 0 | 72.7 |
| Access to care | 7 | 7.3 | 7 | 8 | 0 | 84.8 |

### **Appendix 7 Results of the third round (importance summaries)**

Only outcomes without agreement after the second round were included for rating in the third round. For pharmacist prescribing within a Common Conditions Service, one additional outcome reached consensus in the third round as critically important, re-consultation with other health care providers/other healthcare utilisation, while the remaining six outcomes did not. For other independent prescribing, six of the remaining eight outcomes reached consensus as critically important, cost of care to service providers, adverse events, cost of care to the healthcare system, mortality, healthcare utilisation, and cost of care to patients.

| **Outcomes** | **Median** | **Mean** | **25^th^ Percentile** | **75^th^ Percentile** | **% of rating of 1-3** | **%of rating of 7-9** |
| --- | --- | --- | --- | --- | --- | --- |
| **Common Conditions** | | | | | | |
| Re-consultation with the pharmacist | 6.5 | 6.5 | 6 | 7 | 0 | 50 |
| Re-consultation with other health care providers/other healthcare utilisation | 7 | 6.9 | 7 | 7 | 0 | 76.7 |
| Prescribing rate | 6.5 | 6.7 | 6 | 7 | 0 | 50 |
| Adverse events | 7 | 6.7 | 6 | 7 | 3.3 | 70 |
| Cost of care to patients | 7 | 6.7 | 6 | 7 | 0 | 63.3 |
| Cost of care to providers | 6 | 6.2 | 6 | 6 | 0 | 23.3 |
| GP workload | 6 | 5.8 | 6 | 6 | 3.3 | 16.7 |
| **Independent Prescribing** | | | | | | |
| Mortality | 7 | 6.8 | 7 | 7 | 0 | 76.7 |
| Healthcare utilisation | 7 | 6.9 | 7 | 7 | 0 | 76.7 |
| Prescribing pattern (rates and changes) | 7 | 6.9 | 6 | 7 | 0 | 66.7 |
| Adverse events | 7 | 6.9 | 7 | 7 | 3.3 | 86.7 |
| Quality of life | 6 | 5.7 | 5 | 7 | 6.7 | 33.3 |
| Cost of care to patients | 7 | 6.9 | 7 | 7 | 0 | 76.7 |
| Cost of care to service providers | 7 | 7.1 | 7 | 7 | 0 | 90 |
| Cost of care to the healthcare system | 7 | 7.3 | 7 | 8 | 0 | 86.7 |

### **Appendix 8 Changes in the percentage of critically important ratings across Delphi rounds**

As illustrated, a notable shift in the percentage of experts rating outcomes as critically important was observed in several outcomes (see Figure 7 below). For common conditions, outcomes such as symptom resolution, referral to or re-consultation with other healthcare, cost to healthcare system and access to care were rated below the 75% threshold for inclusion in earlier round (s) and surpassed this threshold by subsequent round (s). A similar trend was observed for some outcomes for evaluation independent pharmacist prescribing outside the context of common condition services. These included mortality, clinical effectiveness, healthcare utilisation, adverse events, patient experience, economic outcomes and access to care.

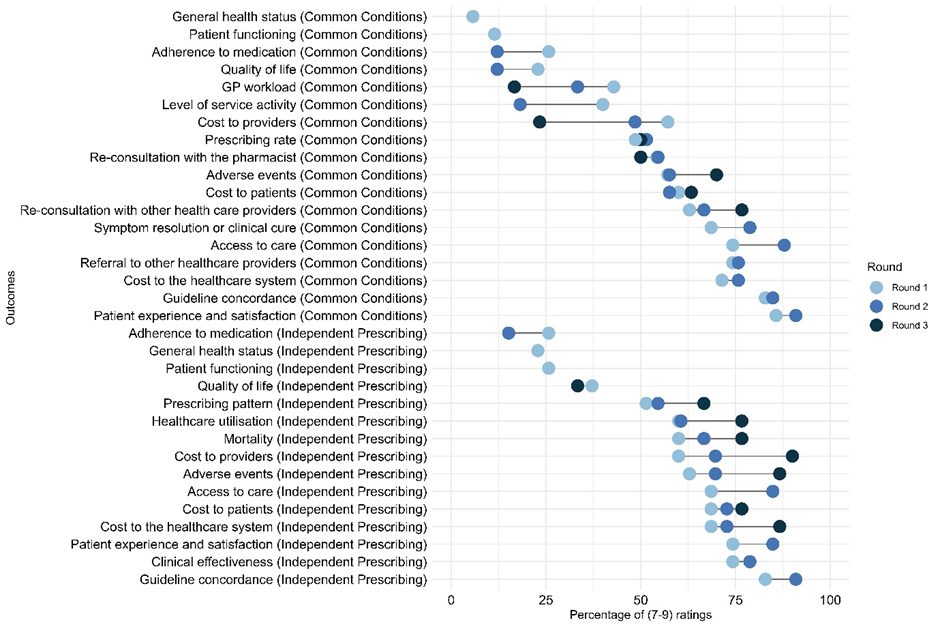

Figure 7 Changes in the percentage of critically importance ratings (7-9) across Delphi rounds

### **Appendix 9 Summary of experts’ qualitative feedback relating to outcome importance**

| **Outcome (description)** | **Qualitative feedback summary**  **(⊕ positive perspective & ⊖ sceptical perspective** | **Examples of representative quotations** |
| --- | --- | --- |
| **Common Conditions Service** | | |
| Symptom resolution or improvement, or clinical cure  i.e. resolution of the condition/symptoms, or positive changes in symptoms from baseline in terms of severity, frequency and/or duration. | ⊕ Most experts highlighted this as an indicator of appropriate diagnosis, clinical judgement and prescribing. It was highlighted as the key aim of prescribing for an ailment, and pharmacist prescribing should not lead to poorer outcomes or delay in accessing effective treatment.  ⊖ Some noted challenges in routinely gathering this data (noting it may require specific follow-up), that resolution/deterioration depends on other factors as well as treatment and may resolve naturally, and the necessity to have an appropriate comparator (e.g. GP data). | *“Clinical cure/symptom resolution or improvement would be an indicator of the selection of the appropriate strategy by the pharmacist i.e. prescribe under the Common Conditions or refer onwards”*  *“...it will be challenging to get this information as routine. This information is not routinely gathered for other prescribers. Follow up of a select number of pharmacist consultations for clinical cure or resolution of symptoms should be undertaken”* |
| Re-consultation with the pharmacist  i.e. returning to the pharmacist for the same condition, related symptoms, or adverse effects after the initial consultation. | ⊕ Most experts highlighted that re-consultation can indicate initial diagnosis and treatment were effective, if onward referral is needed, and could support learning about adapting/expanding scheme. It was noted that re-consultation may be appropriate in some cases, and that the reason for re-consultation is important (e.g. same episode, versus occurrence of a new episode of the condition). The ease of re-consultation where needed (e.g. for unresolved symptoms, possible adverse effects) and potential for continuity of care were seen as positives.  ⊖ Some noted that as re-consultation can be due to many factors (e.g. antimicrobial resistance, not taking treatment as prescribed, a more complicated diagnosis than suspected), it may not reflect quality/appropriateness of care. One expert noted that with appropriate safety netting and advice on seeking further medical care, re-consultation is unlikely to add much to care. | *“Re-consultation with the pharmacist may alert the pharmacist to a need for onward referral (more complex condition than originally thought, possible underlying condition) and possible refresh of learning needs if onward referral may have been the most appropriate strategy originally”*  *“Re-consultation could be for a number of reasons and not necessarily an indicator of quality.”* |
| Referral to other healthcare providers  i.e. the pharmacist refers the patient back to another healthcare provider for further evaluation or treatment. | ⊕ Most experts highlighted that referrals demonstrate adherence to scope of practice and awareness of limitations, and are appropriate in various scenarios (e.g. patient/symptoms are outside scope, uncertainty about diagnosis/treatment, escalation if treatment not effective, symptoms deteriorate or adverse drug reaction occurs). Some noted its importance as an aspect of cost-effectiveness and resource implications (either freeing up or increasing burden on other healthcare professionals’ time).  ⊖ Some cited concerns that referrals may be appropriate or inappropriate depending on the circumstances, the outcome may be hard to interpret given confounding factors, and lack of formal pathways for pharmacists to refer to other healthcare professionals currently. | *“Important to demonstrate adherence to scope of practice and awareness of limitations, which are important aspects of the governance of good prescribing”*  *“…but find it hard to see how it can be translated to a positive or negative impact given the number of potentially confounding factors”* |
| Re-consultation with other health care providers/other healthcare utilisation  i.e. seeking care from other healthcare providers or utilising other healthcare services (e.g. emergency department visits or hospitalisations) after the initial consultation or follow-up visits with the pharmacist relating to the same condition/symptoms due to, for example, treatment failure, unresolved symptoms, or prescribing-related issues. | ⊕ Experts highlighted that such re-consultations are important in instances of unresolved/worsening symptoms, prescription-related side effects or conditions outside the agreed scope of practice. A few highlighted it as important where the patient may not return to the pharmacy but has had to seek additional treatment/advice. Some highlighted its role in evaluating cost effectiveness and assisting pharmacists’ future learning and development and role expansion.  ⊖ Others questioned its significance as an indicator of whether symptoms improved or not, and that it would be complex and not practical to measure using current healthcare infrastructure. Others noted that it may not be important for conditions that are self-limiting, its concept overlaps with previous outcomes, that such an outcome is not assessed for other professionals’ prescribing, or considered it as a process measure rather than measuring impact. | *“Additionally, having a clear referral pathway for prescribing pharmacists will ensure the patient receives the highest standards of care while documenting outcomes for future learning and scope of practice”*  *“I do not think this will add anything in treating these common conditions especially as most are self-limiting”* |
| Prescribing rate  i.e. prescribing rates at level of individual pharmacists or all pharmacists relative to overall prescribing rates of specific medicines or drug classes (e.g. antimicrobial agent for UTI, corticosteroid nasal spray for allergic rhinitis). | ⊕ Many experts considered it a key audit measure or performance indicator of the appropriateness of service delivery and the selection of the appropriate strategy by the pharmacist (either to treat or refer). It could indicate aggregate impact on medicines utilisation nationally and allow comparisons of pharmacist and other prescribers. Others emphasised its importance for some drug classes particularly (e.g. antimicrobials), and to allow comparison with other prescribers. Some noted it would be useful at the level of individual pharmacists, to understand their prescribing relative to peers, and as an indication of frequency of management and therefore familiarity with conditions, likely leading to more effective prescribing.  ⊖ Others questioned whether it provided any indication of quality/appropriateness of care on its own, and numbers would need to be interpreted cautiously alongside referrals and other outcomes, and considering the context such as pharmacy location, patient volume and local outbreaks. Others noted some patients will still prefer to consult with others e.g. their GP, that commercial considerations could drive prescribing, and had concerns about measurement challenges, including the need for a comparator and an appropriate denominator. | *“Prescribing rate could possible be an indicator of the selection of the appropriate strategy by the pharmacist i.e. treat or refer”*  *“…pharmacist participation in service provision would be represented in this number however, limited by the fact that referrals will not be captured”* |
| Guideline concordance/appropriateness of medications  i.e. pharmacists' treatment following clinical guidelines or protocols, and the appropriateness of prescribing (e.g. appropriate drug selection, dose and frequency), acknowledging the role of individual patient’s goal of therapy, preferences, etc. | ⊕ Most experts emphasised its importance to measure evidence-based practice and support patient safety and optimal outcomes. Some noted it would allow comparison to other healthcare professionals to ensure comparable standards of care (while not holding pharmacists to a higher bar than others). Others highlighted that the existence of guidelines/protocols would support pharmacist development and confidence.  ⊖ Some raised concerns about the challenges of measurement, who would assess appropriateness, and current system limitations (e.g. pharmacists’ access to blood results to assess appropriateness). They also stressed that deviations from protocols or prescribing guidelines may be appropriate in some circumstances based on individualising care, and should be documented, clinically justified, and subject to audit. Some suggested it was more important to focus on correct diagnosis (as appropriate treatment will easily follow from this) or protocol compliance i.e. giving advice, over-the-counter medications or referring in correct circumstances. | *“Guideline concordance/appropriateness of medications is important as an indicator that pharmacists recognise adherence to guidelines as the way to ensure the expected patient outcomes”*  *“Notably, consideration must be given to the fact that the current community pharmacy model does not routinely provide pharmacists with access to blood results so use of such a measures to assess impact needs to reflect same. For example, prescribing of an antiviral agent for shingles needs clarification of renal function to enable the pharmacist to select the correct dose and frequency. Factoring in available resources on which to make prescribing decisions would be, in my opinion, an important aspect of such an assessment”* |
| Patient adherence to medication  i.e. how well a patient’s behaviour aligns with the prescribed instructions for taking medication to achieve therapeutic objectives. | ⊕ Some experts emphasised its importance as a factor that will influence outcomes and may explain cases of treatment failure if re-consultation occurs, and that it could reflect the quality of communication practice in pharmacy and potentially highlight issues to be addressed (e.g. further patient education, side effects occurring which lead to non-adherence).  ⊖ Many questioned the ability to accurately measure it or to establish any causal relationship with the prescriber behaviour, e.g. due to lack of comparator and its multifactorial nature (e.g. patient factors). Others questioned the importance in the context of common conditions as short term acute problems. | *“Although I believe this is an important issue, I don't believe it is a reasonable outcome measure for measuring the impact of pharmacist prescribing and is contingent on many other variables”*  *“Patient adherence is a multi-factorial outcome; any causal relationship with the prescriber behaviour would be difficult to establish. Moreover, I don't* [think] *this would be a standard commonly considered in assessing the performance of GP prescribing, so unclear why it would be applied to pharmacists”* |
| Adverse events  i.e. adverse clinical outcomes associated with pharmacist prescribed medications (e.g. drug related adverse effects such as stomach upset) or the consultation (e.g. medications allergy after not confirming allergies). | ⊕ Experts emphasised the importance in ensuring patient safety, accurate diagnosis, appropriate treatment, and continuous pharmacist learning/improvement. While they acknowledged that medication side effects are often unpredictable and can occur regardless of setting/prescriber, they underscored the need to capture all adverse effects, whether resulting from the medication or the service. It was noted that wider access to treatment could identify new adverse effects.  ⊖ Experts noted challenges in accurately measuring this outcome, such as the lack of a comparator, difficulties in collecting relevant data, and the multifactorial nature of side effects. They also pointed out concerns over defining adverse events and assessing the clinical significance, and that they may only be relevant if directly linked to inappropriate prescribing. | *“For me, this is important because of patients' safety, accuracy of diagnosis, appropriateness of treatment, and the potential for more people using these medicines.”*  *“Important yet influenced by many other factors”* |
| Patient experience and satisfaction with care  i.e. experience and satisfaction overall, or with aspects such as access to care (e.g. easiness and convenience), interpersonal communication (e.g. shared decision making), continuity and coordination, comprehensiveness of services, and trust in healthcare providers. | ⊕ Most experts emphasised the importance of capturing the patient perspective, acceptability, public trust, and service user feedback for evaluating accessibility, timeliness, affordability, effective communication, shared decision-making, and the continuity and sustainability of care.  ⊖ Some experts highlighted measurement and interpretation challenges, noting the lack of comparator and the subjective nature of patient satisfaction—particularly when high-quality care involves not prescribing medication. Some highlighted that the lack of measurement in other settings makes it interesting but not critically important. | *“Such data may be important to support the continued expansion of the role and reinforce the trust the community place in the pharmacy profession”*  *“Important but of limited value; sometimes good care is not prescribing and the patient may be unhappy”* |
| Quality of life  i.e. perceived quality of a person’s daily life, assessing their well-being or lack thereof. This includes all emotional, social, cognitive and physical aspects of the individual's life, and may focus on how a person’s well-being and perceived general health is impacted by a condition or disease, and their ability to perform usual activities (now incorporating general health status and functioning since the first round). | ⊕ Some experts highlighted the importance of considering quality of life in healthcare, a few noted that ease, timeliness and convenience of accessing healthcare and treatment will likely improve quality of life and general health status.  ⊖ Many others questioned the important due to the multifactorial nature and potential for confounding factors, felt that prescribing for common conditions is unlikely to impact quality of life, needing long follow-up to identify any impact. Others noted that patient experience and satisfaction was a more important outcome. | *“Pharmacist prescribing can improve quality of life by providing easier access to appropriate care for common conditions. People may not need to take time out of work and may also be able to access treatment for their condition in a more timely manner. This can lead to improved well-being as some of the stress of accessing healthcare has been removed. Some people will also find it easier to approach a pharmacist about their condition and there receive more timely interventions”*  *“Although I believe this is an important issue, I don't believe it is a reasonable outcome measure for measuring the impact of pharmacist prescribing and is contingent on many other variables”* |
| Cost of care to patients  i.e. direct and indirect costs to patients of accessing pharmacist prescribing, on its own, relative to alternatives, or relative to the time to access care. | ⊕ Experts emphasised the importance to assess the overall impact of the service, as it may influence equitable access and treatment adherence. Some noted its role in informing future audits or government funding decisions. Some highlighted the importance of evaluating cost-effectiveness in the context of time to access care, compared to alternative care pathways.  ⊖ Many other questioned it as a measure of impact, and some felt clinical appropriateness was more important. Some felt the importance would depend on whether the service is funded by the government/health service. The challenges of using this to assess impact (e.g. necessity of comparator data, challenges with indirect cost measurements, circumstances that may influence such as medicine shortage) were noted, and a perception of pharmacists’ conflict of interest as commercial entities (though this was perceived to be minimal). | *“Cost of medical consultation and treatment is vital as it can be a reason for non-compliance with treatment”*  *“It depends on whether these services will be funded or not. If the cost is higher than seeing a GP then it will not be perceived as good value from a monetary perspective.”* |
| Cost of care to providers  i.e. cost-effectiveness and cost minimisation (considering direct and indirect costs) from the perspective of community pharmacies providing the common condition service. | ⊕ Experts emphasised its importance to measure cost-effectiveness and determine availability, sustainability and accessibility of the service, especially from the perspective of community pharmacies as service providers. It was noted that such costs may restrict number of providers if prohibitive, and this service will take pharmacists away from other roles.  ⊖ Other noted that it was less important as it is a part of cost to the health system overall, and any assessment should also consider any cost savings across the system. It was also noted that it would be difficult to capture ancillary activities that often happen during GP consultations for minor ailments (e.g. identifying another issue, chronic disease screening). | *“Useful to determine overall costs to providers to determine sustainability of the programme from the pharmacist's point of view”*  *“Cost of care to providers, I believe, is part of the wider healthcare system infrastructure cost so considering assessment of costs for the patient, providers and the healthcare system need to be balanced with cost savings. For example, early intervention in the treatment of shingles and associated costs vs. savings in terms of admission to hospital and bed days.”* |
| Cost of care to the healthcare system  i.e. cost-effectiveness and cost minimisation (considering direct and indirect costs) from the perspective of the healthcare system. | ⊕ Many experts emphasised its importance as a benefit to the whole healthcare system, and that the service should only continue if it is a cost-effective means of providing care, considering direct, indirect and opportunity costs. Some noted that assessing potential cost saving would be important, as this may support future funding/expansion.  ⊖ Other noted costs not being a major issue for common conditions, highlighting challenges with capturing costs particularly any downstream effects of the service, and the need to balance cost with quality of care. | *“Understanding the cost-effectiveness of this scheme is important; it should not be pursed if there are more cost-effective ways of providing the same level of patient care”*  *“Cost not a major issue in the management of common conditions”* |
| GP workload  i.e. increase or decrease in number or length of consultations with general practice staff  due to task shifting of common conditions consultation and/or additional referrals or visits. | ⊕ Experts emphasised its importance given the current GP shortage and inability to get appointments with some GPs. Some noted the importance given that some of the rationale for the service is to reduce the workload of general practice staff and also to ensure it does not increase GP workload.  ⊖ Others questioned whether any reduction in GP workload would be expected, as GPs may divert capacity to manage more complex cases. Others noted challenges of controlling for confounding factors and that increasing supply (by introducing pharmacist prescribing) may increase demand. | *“Would free up time for GPs to deal with other matters”*  *“Given that this is one of the drivers for introducing the common ailments scheme, it would seem like a sensible surrogate measure - however, the ambition is that the capacity freed up by treating minor conditions will be diverted to managing more complex cases, and therefore I would not anticipate that there would be any reduction in GP workload and I therefore question the usefulness of the measure”* |
| Access to care  i.e. measures such as the proportion of eligible people having a consultation or receiving medicines, the number of overall dispensed prescriptions, and time to receipt of prescriptions, equity of access and perceived convenience of accessing care. | ⊕ Experts emphasised its importance as a major goal is to provide wider and more convenient access to care in a timely manner. This was noted as particularly important given current limited capacity in the health system, and in aligning with Sláintecare aims.  ⊖ Other questioned the examples given in the description such as the number of overall dispensed prescriptions, and noted that some aspects of access, including equity, may be more important than others. | *“One of the drivers of the scheme is to increase access to patients, so this seems an important outcome measure”*  *“Perceived convenience of care will be important and will affect the level of service use, not sure about the other examples given”* |
| Level of service activity  i.e. overall volume of patients availing of the service, number of prescriptions issues, or the numbers of service claims. | ⊕ Experts emphasised its importance as a measure of pharmacists’ workload, and to inform future changes.  ⊖ Experts also noted the necessity to assess it across a long period and comparatively with other states and areas, and that as a measure of quantity was less important than quality. Expected variation between, for example, pharmacies in a city versus a village, was also noted. Others were unsure what this means or felt that itis covered by other outcomes. The impact prescribing activity may have on workload and time patients may wait to have medications dispensed was also noted. | *“Determining workload of pharmacists, informing future changes for regulation of pharmacy in the future”*  *“It may take time for the level of activity to increase to a stable level but it should be reviewed in comparison with the service in other states”* |
| **Other Independent Prescribing** | | |
| Mortality  i.e. death from any cause or specific causes. | ⊕Some experts emphasised its importance to measure in the event of sudden deterioration or prescribing-related fatal adverse effect. A few noted there could be improvement in this outcome linked to pharmacist prescribing, and that it should be part of standard outcome analysis and mortality review where already in place. Its importance as a public health measure, and providing an overall focus on accountability and safety was also noted.  ⊖ Many questioned the ability to accurately measure it or to establish causality with independent prescribing outside of a controlled trial. Some felt medication-related mortality may have some relevance but less so overall mortality. It was also noted that where pharmacist prescribers worked as part of a team it may be less important in measuring the impact, but may be more important in other contexts. | *“I see this as applicable to all prescribers where it must be contextual (i.e. drug related or condition related) but considered to ensure patient safety and benefit are maintained as central to all prescribing decisions for pharmacists, as it is applicable to all healthcare professional prescribers. This supports creating a culture of accountability, raising awareness of ADRs* [adverse drug reactions]*, medication related harm and also ensuring independent responsibility for the independent prescribing model”*  *“It would be difficult to establish causality when examining the impact of pharmacist prescribing, apart form in an adverse sense (i.e. the prescribing action led to a fatal adverse drug reaction). Maybe in a controlled sense, where you had one group who did not receive the independent prescribing intervention and one group who did and see the impact of independent prescribing on mortality but the results are likely to be confounded by many other variables”* |
| Clinical effectiveness  i.e. measures of the specific benefits of prescribed treatments, such as improvement/control of blood pressure, blood glucose and blood cholesterol, reduction in major cardiovascular event risk, prevention of unplanned pregnancies, HIV prophylaxis, etc. | ⊕ Experts emphasised its importance as being central to the intention of introducing pharmacist prescribing. Some noted measures of clinical effectiveness should be equivalent to those used for other prescribers.  ⊖ Others questioned the ability to accurately measure it given complexity of conditions, and confounding factors that may influence clinical effectiveness that would need to be accounted for. Others questioned whether the examples given were patient-centred. | *“Would be great to have robust metrics such as improvements in blood pressure, blood glucose and blood cholesterol”*  *“Clinical effectiveness is not always easy to evaluate: the examples given are very didactic in nature and ignore the patient perspective/values (paternalistic approach)”* |
| Healthcare utilisation  i.e. increased or decreased primary care visits, emergency department visits, or hospitalisations. | ⊕ Experts emphasised its importance to measure effectiveness of pharmacist prescribing, and that it would provide information on potential benefits to patient and the overall healthcare system, or potential increased burden.  ⊖ Others noted that there may not be a major impact on this, that the role of pharmacist prescribers in multiple settings (primary/secondary care) would need to be carefully considered in any analysis, and that assessment is complex (e.g. need for a suitable follow-up period and complete data). | *“Benefits to the patient and to the overall healthcare system should be the most important factors in making a decision on these proposals and a measure of healthcare utilisation will provide essential information in this regard”*  *“Important to consider but may not have a major impact in the grand scale”* |
| Prescribing pattern (rates and changes)  i.e. rates of prescribing of specific medicines or drug classes, changes made to prescribed treatment, or initiation or discontinuation of medicines. | ⊕ Experts considered it important to audit and assess independent pharmacist prescribing, and could capture reductions in overprescribing medicines. It was felt measurement of this outcome should be the same as for other prescribers. It was also noted as important to inform considerations about supply chain or identify potential overuse of some medicines e.g. antimicrobials.  ⊖ Experts expressed concerns about measurement methods, and potential changes in medicines utilisation (e.g. potential for misuse, reduction in over the counter sales). Others also highlighted the need for a system that integrates with other prescribers to manage patient complexities (e.g. prescribing for one condition in the context of multimorbidity), and to focus on the quality of prescribing and drug selection. | *“Reduction in prescribing of commonly over prescribed drug classes would be a welcome change. Pharmacists well placed to optimise medicines and deprescribe long term medicines for which potential harms are outweighing potential benefits”*  *“The pharmacist needs a system to collaborate with the GP around prescribing, and ideally would not make recommendations about one disease in isolation: the recommendations must reflect the real-world complexity of patients with multimorbidity & polypharmacy”* |
| Guideline concordance/appropriateness of medications  i.e. pharmacists’ treatment following clinical guidelines or protocols, and the appropriateness of prescribing (e.g. appropriate drug selection, dose and frequency, prescribing omitted medications, discontinuing inappropriate medications). | ⊕ Most experts highlighted the outcome's importance for ensuring adherence to evidence-based practices and delivering safe and effective care. Some noted it would capture the contribution of pharmacists and their ability to tailor medicines, and support public trust.  ⊖ Others emphasised the need to consider the complexities of real-world patient situations beyond merely adhering to the latest guidelines for single diseases, and that it may be appropriate to deviate from guidelines based on individual circumstances (and reasons for this should be captured). | *“I feel it's of utmost importance that all guidelines are followed to ensure proper care & reduce adverse effects/outcomes for users”*  *“Guidelines support good care but are often developed for 'single diseases' while excluding huge swathes of the population (elderly, multi-morbid, palliative etc etc). We need shared decision making with deep patient engagement: not unthinking adherence to the latest single disease guideline”* |
| Patient adherence to medication  i.e. how well a patient’s behaviour aligns with the prescribed instructions for taking medication to achieve therapeutic objectives. | ⊕ Some experts emphasised that appropriate consultation skills and medication choice are important to ensure patient adherence particularly in chronic conditions.  ⊖ Many questioned the ability to accurately measure it, and its importance due to lack of comparator information and the many factors that can influence adherence. | *“Patient adherence to treatment to vital when dealing with chronic conditions”*  *“As mentioned for common conditions, this outcome in my opinion relates not only to prescribing but to other aspects of medication management”* |
| Adverse events  i.e. adverse clinical outcomes associated with both pharmacist-prescribed medications (e.g. hypertension from rapid antihypertensives titration) or the intervention (e.g. chest pain overlooked/misdiagnosed as heartburn). | ⊕ Experts highlighted the outcome's importance for patients' safety and measuring this at the local level could support additional training/learning. It was noted that measuring this outcome would provide further evidence on benefit-risk assessment for medicines.  ⊖ Experts noted measurement challenges, its multifactorial nature and the need to clarify its definition (i.e. drug related or pharmacist consultation related), and that these two categories may have different levels of importance. | *“Those that are a direct result of a pharmacist-prescribed medication or intervention should be recorded and shared for patient safety and additional training/learning purposes. Not to be recorded for punitive reasons.”*  *“Not all adverse effects can be anticipated so it needs to be clarified if it is due to the pharmacist prescriber or to the medication”* |
| Patient experience and satisfaction with care  i.e. experience and satisfaction overall, or with aspects such as access to care (e.g. easiness and convenience), interpersonal communication (e.g. shared decision making), continuity and coordination, comprehensiveness of services, and trust in healthcare providers. | ⊕ Experts highlighted the importance to capture the patient's voice and trust, particularly in the setting of chronic diseases where continuity of care and communication have impacts over a longer period of time.  ⊖ Some experts highlighted that in some cases it may not reflect the actual context of the care provided, particularly with measuring patient satisfaction which is more subjective. | *“I think this is more relevant in the setting of pharmacist prescribing in the setting of chronic conditions outside of common conditions alone where continuity of care and communication have impact over a longer period of time”*  *“This is somewhat subjective so while important, may not be indicative of the actual scenario. Patients have a voice and that must be valued across the entire healthcare system. Positive and negative experiences will be communicated in the community and possibly shared across social media channels”* |
| Quality of life  i.e. perceived quality of a person’s daily life, assessing their well-being or lack thereof. This includes all emotional, social, cognitive and physical aspects of the individual's life, and may focus on how a person’s well-being and perceived general health is impacted by a condition or disease, and their ability to perform usual activities (incorporating general health status and functioning since the first round). | ⊕Some experts emphasised the importance of quality of life in independent prescribing for chronic conditions, where longer follow-up is possible, unlike for common conditions. Some also expressed the importance of considering general health status and functioning when the treatment is intended to exert benefits in improving this outcome, and that person-centred care delivered by pharmacists could also improve it.  ⊖ Many others questioned the importance due to the multifactorial nature and potential for confounding factors, and the limited sensitivity of measures. Others saw measuring these outcomes as low priority, if pharmacist prescribing is unlikely to impact this, especially if it took up pharmacist resource to measure it. | *“I feel this is much more important when dealing with chronic conditions as opposed to minor ailments. Any negative impacts on quality of life are far more compounding for someone with a long term condition in comparison to someone with a limited condition”*  *“Quality of life measures unlikely to be impacted by single episodes of pharmacist prescribing”* |
| Cost of care to patients  i.e. direct and indirect costs to patients of accessing pharmacist prescribing, on its own, relative to alternatives, or relative to the time to access care. | ⊕ Experts emphasised the importance of the overall cost to patients for equitable access and sustainability of the care, particularly for chronic conditions. Some highlighted the importance of collecting related data for transparency and government funding considerations. Some highlighted the importance of evaluating the cost-effectiveness in terms of both money and time and the impact on service uptake.  ⊖ Others questioned cost as a key measure in secondary care and argued its relevance depends on funding models, while some noted that it can be influenced by many factors. | *“I think it's very important to consider cost of care to patients in terms of providing a service that is sustainable and equitable and the potential cost of not being able to access such services in terms of e.g. hospital admissions”*  *“Can be influenced by many factors. Also depends on the specific health service. In some countries there is no charge for prescription medication”* |
| Cost of care to service providers  i.e. cost-effectiveness and cost minimisation (considering direct and indirect costs) from the perspective of institutions providing the care (e.g. community pharmacies, GP practices, hospitals). | ⊕ Experts stressed the importance of overall cost to providers for assessing value, long-term availability and sustainability of the care, noting that cost of implementation of service should be considered. Some noted that collecting related data at national level could help future policy decisions, and to show the value of pharmacist prescribing (e.g. cost saving from medicine optimisation and fewer hospital visits).  ⊖ Some noted that cost should not be more important than service quality, and that the cost might differ across providers (e.g. small independent pharmacies versus pharmacy chains). | *“Again, not an area I'm knowledgeable in, but I'd imagine it pretty critical to enable pharmacists to firstly enable pharmacists to incorporate these services and secondly to make it viable in the long term”*  *“Again disparities may arise when comparing small independent pharmacies and large chain pharmacies like Boots etc”* |
| Cost of care to the healthcare system  i.e. cost-effectiveness and cost minimisation (considering direct and indirect costs) from the perspective of the healthcare system. | ⊕ Many experts emphasised the importance to assess benefit to the whole healthcare system, and to contribute to cost-effectiveness and budget impact in considering time and financial cost. Some noted that assessing potential cost saving (direct and indirect through reduced hospitalisations for example) would be important to show the value of the independent pharmacists prescribing in this setting and support future funding/expansion.  ⊖ Other noted the need to balance cost with quality of care, and that the latter is more important. It was also questioned whether other pharmacist roles may provide greater value and benefit to the healthcare system. | *“...all of costing outcomes are very important if aiming to demonstrate cost-effectiveness and investigate budget impact as part of the project”*  *“Quality of service should way more than cost ideally”* |
| Access to care  i.e. measures such as the proportion of eligible people receiving medicines, the number of overall dispensed prescriptions, time to receipt of prescriptions, equity of access and perceived convenience of accessing care. | ⊕ Experts emphasised its importance as a key rationale for the expansion of pharmacists’ role. It could widen access to those who need but do not currently access care.  ⊖ Others questioned its importance in secondary care compared to primary care, that it should be balanced with quality, and others highlighted the importance of considering equity as part of access. | *“Important as one of the aims is to increase access to care”*  *“Again more likely to be important in primary care rather than secondary care. Secondary care likely to be more specialised prescribers with the potential to be involved in outpatient clinics”* |

### **Appendix 10 Experts’ feedback on feasibility of outcome measurements**

In free-text responses, the Delphi expert panel emphasised that there is no standardised approach in current pharmacy software to record data necessary for measuring these outcomes, meaning pharmacists would have to rely on local documentation systems that cannot accurately and consistently capture all these measurements. The expert panel suggested two main solutions to capture required data for evaluation the impact of pharmacist prescribing in a Common Conditions Service. One interim solution included creating a new digital solution for the Common Conditions Service, similar to the digital solution for the Emergency Contraception Scheme, or a similar legislative record keeping requirements system to the PharmaVax system. The second solution is to adapt the IT infrastructure and care pathways to capture required data. Some experts further suggested that the National ePrescribing System (NePS) being developed by the Health Service Executive (HSE) could be adapted to record any required data, while others proposed modifying existing patient medication record (PMR) software, using standardised coding such as SNOMED and harnessing individual health identifiers (IHI) for each patient once integrated.

Prospective data collection methods, including follow-up visits/calls or patient surveys, were identified as possible interim solutions for capturing clinical and patient-reported outcomes. One Delphi expert gave an example of the Sláintecare Migraine Pilot, which used IPUnet with external survey software, suggesting that prospective data collection or patient follow-up would be necessary for the symptom improvement outcome. Some Delphi experts highlighted that data on re-consultation with the pharmacist could be accurately measured if IHIs were incorporated successfully into pharmacy PMR systems. Other suggested that this data could be reflected in routine payment or as a record-keeping feature added to PMR systems (for example recording such consultations as a product or intervention). Some Delphi experts pointed out that the Healthmail facility (e.g. surgery Healthmail) may support pharmacist-to-GP referrals, but emphasised the need for standardisation, coding, and a common patient identifier, along with SNOMED-coded interventions to facilitate data capture. Experts added that, in the interim, pharmacists could document referrals in their PMR notes or email logs. Experts also highlighted the need for standardised intervention coding and patient identifiers that could facilitate data sharing, to support re-consultation with other healthcare providers, via PMR systems, Shared Care Records (SCR)/My Health App, and GP systems through Healthlink. Moreover, experts added that a referral mechanism similar what GPs have would be useful following the implementation of Shared Care Records within the HSE.

For adverse events, Delphi experts suggested prospective data collection or data from Health Products Regulatory Authority (HPRA) as potential sources. They highlighted that current pharmacy systems lack the capability to systematically capture minor adverse events or non-drug-related outcomes. Some suggested that PMR software modifications or an online portal for pharmacist prescribing (e.g. having a 'modified' version of the National Veterinary Prescription System (NVPS)) could improve adverse events capture. Some experts highlighted that prescribing rates could be captured via PMRs and existing pharmacy dispensing software if modifications were made to track pharmacist prescribing (e.g. pharmacists using their Pharmaceutical Society of Ireland number in the PMR and a tick box if medicines were prescribed by the pharmacist). HSE Primary Care Reimbursement Service (PCRS) reimbursement data was noted as a reliable source if the services are funded and reimbursed, and wholesaler commercial data was suggested as a possible supplementary data source. Others proposed a separate prescribing portal would be required to track prescribing rates among private patients if the Common Conditions Service is not funded or reimbursed. Despite challenges, experts also suggested integrating a standardised digital checklist within PMR systems to capture guideline concordance, particularly for medicines prescribed under protocols. Others pointed out that guideline adherence could be monitored through structured online forms and data collection tools, similar to those used in vaccination programs.

Many Delphi experts highlighted that patient experience and satisfaction could be measured using existing tools such as patient surveys or digital consultation questionnaires. Some experts also proposed integrating these methods into PMR systems, or independent survey links sent after consultations. Others suggested adapting national and other public health surveys such as the Health Information and Quality Authority National Patient Experience Survey, Healthy Ireland Survey, European Health Survey or the OECD Patient-Reported Indicator Surveys (PaRIS) to be modified or extended to gather these for sample of population. For access to care, some experts pointed out that PCRS reimbursement data could provide insights if the service is publicly funded, while data from clinical research service providers like IQVIA could also be of value. Experts also noted that significant new systems (e.g. a dedicated patient portal or structured research audits) or the HSE My Health App may be necessary to effectively capture these outcomes.

Several experts highlighted that economic outcomes might be easier to measure, however modifications to current systems and additional data collection may be needed to provide a comprehensive assessment. Some experts pointed out that cost of care to patients could be tracked using community pharmacy data and PMR systems (with some modifications) and, if the service is funded, PCRS reimbursement claims would be an appropriate source. They added that modifications to existing PCRS reporting systems to track pharmacist prescribing should be considered to assess costs of care to the healthcare system. It was also noted that costs of care to the healthcare system could be assessed as part of Health Technology Assessments (HTA).
